# Can home spirometry and FeNO testing improve asthma diagnosis? – a feasibility study

**DOI:** 10.64898/2026.04.16.26351022

**Authors:** Ran Wang, Alexander Thompson, Miriam Bennett, Angela Simpson, Stephen J Fowler, Hannah J Durrington, Clare S Murray

## Abstract

**Introduction:** Asthma misdiagnosis is common. Despite temporal variability being a hallmark of asthma, diagnosis largely relies on single clinic-based measurements. We evaluated the feasibility and potential diagnostic utility of home spirometry and fractional exhaled nitric oxide (FeNO) testing.

**Methods:** Symptomatic, untreated adults with GP-suspected asthma underwent routine diagnostic tests including bronchodilator reversibility, in-clinic FeNO, blood eosinophil counts and bronchial challenge testing (BCT). Participants also performed home spirometry and FeNO measurements over two weeks. The reference standard (asthma/not-asthma) was determined by an expert panel of at least two asthma specialists based on clinical history and routine test results; home spirometry and home FeNO measurements (index tests) were blinded to the panel.

**Results:** Of 51 recruited participants, 38 had asthma confirmed or excluded by the panel. Home measurements yielded 1058 spirometry recordings from 37 (73%) participants and 848 FeNO recordings from 39 (76%). Among those completing any home measurement, median (IQR) adherence was 66.7(58.6-97.6)% for home spirometry and 78.5(51.8–103.6)% for home FeNO. In participants with a diagnostic outcome (home FeNO: n=32; home spirometry: n=28), the putative home-testing metrics demonstrated high sensitivities at ≥90% specificity, outperformed diurnal peak expiratory flow. Incorporating home testing into the BTS/NICE/SIGN 2024 diagnostic pathway could reduce reliance on BCT by 65%. Collection of health impact data was feasible, with economic scoping indicating the potential value for money.

**Conclusions:** Home spirometry and FeNO testing for diagnosing asthma were feasible and showed potential to improve asthma diagnosis. These findings support further evaluation through a fully powered diagnostic accuracy study.

## INTRODUCTION

Asthma is the most common chronic airways disease worldwide^1^, but misdiagnosis rate is substantial, with up to 1 in 3 people treated for asthma do not have the disease, while many others with asthma symptoms remain undiagnosed^2–7^. Diagnostic delays and misdiagnosis lead to avoidable patient harm and place a significant burden on healthcare systems^5–7^. In contrast, early and accurate diagnosis followed by appropriate treatment, can reduce unscheduled healthcare utilisation and improve disease outcomes^8^.

The definition of asthma highlights its intrinsic nature of temporal variability^9^. However, the current clinic-based diagnostic approach relies on one-off time-point assessments, regardless of whether individuals are experiencing their usual symptoms at the time^7,9^.

Mounting evidence suggests that asthma exhibits marked temporal variations in disease pathophysiology, with timing of test measurements influencing diagnostic outcome and clinical decision-making^10–15^. While the Global Initiative for Asthma 2026 strategic report recognises the impact of timing of diagnostic measurements and recommends performing key objective testing when patients are symptomatic or in the morning^9^, achieving this in routine care requires flexible and rapid access that is rarely available in practice. Peak expiratory flow diurnal monitoring is currently the only home-based test for asthma; however, its poor diagnostic yield limits clinical utility, and is only recommended when spirometry is not available^9,16^.

Advances in technology now allow tests previously restricted to clinical settings to be performed at home using portable devices, allowing flexible timing of measurements and better capture of variability in airflow limitation and airway inflammation^14^. Whilst early stakeholder evaluations suggest that undertaking home spirometry and fractional exhaled nitric oxide (FeNO) testing at the diagnostic setting are generally welcomed by both patients and healthcare professionals ^17,18^, their clinical feasibility and potential to improve diagnostic accuracy remains uncertain.

This study aimed to evaluate the feasibility and potential diagnostic utility of home-based spirometry and FeNO testing for asthma diagnosis.

## METHOD

### Study procedures

The study is nested in the Rapid Asthma Diagnostics for Asthma study (RADicA, https://www.radica.org.uk, ISRCTN 11676160)^19^. Briefly, symptomatic and untreated adults (≥16 years with no inhaled or systemic glucocorticosteroid use for at least 4 weeks) with GP-suspected asthma were recruited from primary care. Clinical history was taken and physical examination carried out. All participants had routine asthma diagnostic tests performed, including FeNO (NIOX VERO, Circassia, UK, and NOBreath, Bedfont, UK) measured in clinic, spirometry and bronchodilator reversibility (BDR) and bronchial challenge testing (BCT, mannitol and/or methacholine). All participants were skin prick tested for eight common inhaled allergens, blood eosinophil counts measured, and 5-item asthma control questionnaire (ACQ-5) assessed^20^. A trial of inhaled steroids (ICS, fluticasone propionate 250mcg twice daily) was given for 4-8 weeks before diagnostic tests repeated (Figure 1).

**Figure 1.**
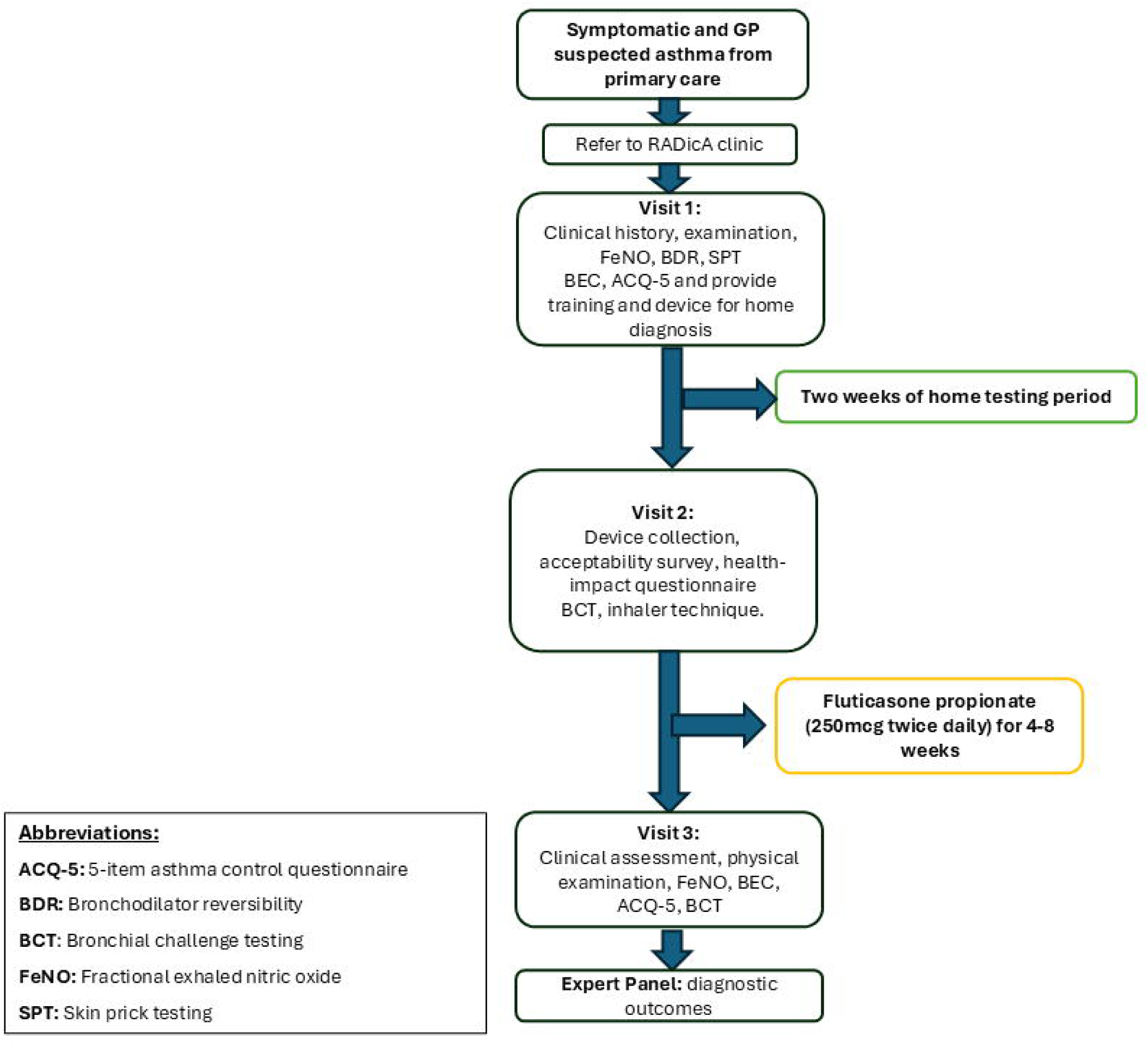
Study flow chart

#### Home testing

Prior to initiation of ICS, all participants were provided with home spirometry (MIR Spirobank Smart, Intermedical, UK) and FeNO (NOBreath, Bedfont, UK) devices. With permission, the mobile software application (‘app’, MIR Spiro Software) associated with the spirometers was downloaded, set up and installed on the participants’ smartphones in the RADicA clinic during their appointments. The MIR Spiro software is compliant with the international ATS/ERS guidelines^21^ and standards for spirometry. The app software also provides a virtual assistant for users to achieve optimum spirometry blow technique. In-person training and written instructions were provided for both devices; the time taken for training for each test were recorded.

Participants were asked to perform FeNO and spirometry tests 4 times a day at home, e.g. 4-5 hourly (±1hr) when awake (e.g. 0600-0800, 1100-1300, 1600-1800, 2100-2400hr) and whenever symptomatic for one week. During the second week, participants were asked to use the spirometer twice-daily (0600-0800 and 2100-2400hr), and to use both spirometry and FeNO whenever they have symptoms. As diurnal PEF monitoring over two weeks remains a recognised diagnostic parameter^9,16,22^, this schedule is designed to mirror established PEF protocol and to evaluate whether home spirometry and FeNO could feasibly capture longitudinal variability within a similar timeframe and test frequency. The four-times-daily schedule in the first week was to enable characterisation of diurnal patterns in test parameters and identification of optimal time points for future refinement.

Participants were told to measure FeNO prior to spirometry measurements, and prior to reliever use whenever possible (and repeat 15-20 minutes after if the reliever was taken). There were no dietary or specific lifestyle changes advised. Visual Analogue Scale (VAS) were used to self-rate symptom burden at time of testing using diary book (Figure E1). All participants were asked to complete a test acceptability questionnaire following the testing period.

Health-economic parameters were collected at Visit 2 using participants administered questionnaire; EQ-5D-5L^23^ is assessed daily during home testing period with questionnaire embedded within the diary booklet.

Symptoms, EQ-5D-5L, and home FeNO measurements were recorded in a diary booklet, while home spirometry reports were collected via the app and emailed by participants after each test.

#### Index tests, reference standard and blinding

The reference standard was established by an expert panel comprising at least two asthma specialists (from SJF, CSM and AS), who confirmed or refuted an asthma diagnosis using all available clinical and routine test data (Figure 1) ^19^. Home spirometry (except for PEF diurnal variability) and FeNO (index tests) results were blinded to panel members.

The study protocol was approved by the North West-Greater Manchester East Research Ethics Committee (18/NW/0777); all participants gave written informed consent.

#### Patient and public involvement

A Patient Advisory Group (PAG) comprising five members with asthma or caring for someone with asthma. The PAG helped design and review all relevant patient-facing materials, including the diary (Figure E1), device-training materials, participant information sheets and consent forms. One member also served on the study’s trial steering committee.

#### Outcomes

The primary outcomes were recruitment and retention rates and test adherence; key secondary outcomes included equipment loss/damaged/failure rate, proportion of patients diagnosed with asthma by the joint British Thoracic Society, National Institute for Health and Care Excellence and the Scottish Intercollegiate Guidelines Network 2024 (BTS/NICE/SIGN 2024) pathway^16^ and by expert panel, putative metrics in asthma and non-asthma and their discriminative ability against reference standard and the feasibility of collecting data for health-economics evaluation.

#### Sample size

A sample size of 50 participants was considered sufficient for a feasibility study, enabling estimation of primary feasibility outcomes while providing preliminary diagnostic performance estimates to inform future diagnostic accuracy study^24^.

#### Statistical analysis

Feasibility data were summarised using descriptive statistics. Adherence rate was estimated based on the proportion of expected measurements successfully received. All FeNO measurements were considered valid and included in subsequent exploratory analyses. For home spirometry, only measurements meeting a minimum quality threshold of grade C or higher based on forced expiratory flow within in 1 second (FEV_1_) were included^24^, unless otherwise stated.

For home-based longitudinal data, multilevel intraclass correlation coefficients (ICC) were used to quantify variance attributable to between-participant, day-to-day, and within-day differences, estimated using mixed-effects models (lme4 and performance R packages). Repeated-measures correlation was used to quantify within-participant associations between objective measures and symptom scores, accounting for repeated observations (rmcorr R package) and were further examined using linear mixed-effects regression models (lme4 packages). Non-parametric data were log-transformed prior to analysis. Diurnal variability of peak expiratory flow (PEF) over 2 weeks were measured using home spirometry and calculated using formula: ∑[(Maximum-minimum)/mean]/(number of days) x 100.

##### Putative home diagnostic metrics

Given that asthma is characterised by variable airflow limitation^10^, putative spirometric indices reflecting within-individual variability in airflow obstruction (e.g. in FEV_1_ % predicted and PEF) were evaluated. For home FeNO testing, both average and variability-based parameters were examined, given their reported associations with relevant clinical outcomes^10,25^.

Diagnostic performances of the putative home testing parameters were assessed using receiver operating characteristic area under the curve (AUROC) analysis (pROC package). As no single asthma diagnostic tests is sufficient to rule out asthma, the national and international diagnostic guidelines adopt sequential rule-in approaches^7,9,16,22^ and high specificity tests (low false positive rate) are prioritied^16^. We report putative diagnostic thresholds and the corresponding sensitivity estimates achieving ≥90% specificity.

Confidence intervals for AUC, sensitivity, and specificity were derived using bootstrap resampling with n=1000 iterations.

All analyses were conducted using R (version 4.5.1) within RStudio (Version 2023.12.1).

## RESULTS

### Feasibility

#### Study feasibility and test adherence

Of 67 eligible participants, 51 (76%) consented to participate in the sub-study. Two-thirds (63%) were recruited from areas in the most deprived tertile of the Index of Multiple Deprivation. There was no difference in baseline characteristics between those who consented and declined (Table 1). Of the 51, definitive diagnosis could be confirmed by the expert panel in 38 participants (25 [65.8%] of whom had asthma) (Table E1). Following the current BTS/NICE/SIGN 2024 guidance^10^, 42 participants had confirmed diagnostic outcome, of whom 66.7% had asthma (Table E2). Agreement between BTS/NICE/SIGN 2024 and expert panel enhanced reference standard was 78.4%.

**Table 1.** Baseline characteristics table in those who consented and declined participation in home testing.

|  | **Declined**<br>**(n=16)** | **Consented**<br>**(n=51)** | ***p-value*** |
| --- | --- | --- | --- |
| Male, n (%) | 6 (37.5%) | 21 (41.2%) | 1 |
| Age, yrs, median (IQR) | 32.5 (22.8- 39.5) | 30 (21.0, 39.0) | 0.837 |
| Ethnicity, white, n (%) | 7 (43.8%) | 28 (56.0%) | 0.832 |
| Index of Multiple Deprivation (decile<sub>≤</sub>3), n (%) | 9 (56.3%) | 32 (62.7%) | 0.864 |
| Body mass index, kg/m<sup>2</sup> | 28.0 (24.3-32.2) | 28.1 (24.1-35.2) | 0.686 |
| Shift worker, n (%) | 6 (37.5%) | 7 (14.0%) | 0.083 |
| ACQ points at presentation | 1.6 (0.8-2.4) | 1.4 (0.6-2.2) | 0.636 |
| Lost in follow up* | 2 (12.5%) | 8 (15.6%) | 1 |
| % asthma diagnosis | 10 (62.5%) | 25 (49.0%) | 0.665 |

\*Did not attend at visit 2

Whilst 1058 home spirometry measurements were obtained from 37 (72.5%) participants, of which 602 (56.9 %) were of adequate quality (grade C or above), 848 home FeNO readings were obtained from 39 (76%) participants (Figure E2-3). Of those who performed at least one home measurement, the median (IQR) adherence rate for spirometry was 66.7 (58.6-97.6) % and FeNO 78.5 (51.8-103.6) %.

There were no devices lost or damaged. One participant experienced difficulty using the spirometry app during an app update period. In three participants, non-attendance at the follow up visit 2 necessitated retrieval of the device by travelling to the participant’s home. The median (IQR) training time was 9.0 (6.8-11.0) minutes for home spirometry and 5.0 (2.5-7.0) minutes for FeNO. Home testing strategy using both tests were acceptable in most participants (Figure E4-5).

#### Feasibility of health economics data collection

35 participants (68.6%) completed at least one daily EQ-5D-5L assessment at home, providing 307 participant-days of utility data. Completion rates were similar between participants with and without asthma (Figure E6-7, Table E3). Health impact questionnaires were completed by 37 participants (72.5%; 33 with a diagnostic outcome); completion rates for health impact measures, productivity data and out-of-pocket costs all exceeded 70%.

30% (11/37) of participants reported at least one unscheduled healthcare utilization due to respiratory symptoms within the previous 3 months (Figure E8).

### Exploratory analyses

#### Longitudinal variability

The variability observed in subjective symptoms, and objective home FeNO and spirometry measurements was substantial (Figure 2, Figures E9-E16). 18 (46.2%) participants had home FeNO readings that straddled the diagnostic cut-off of 50ppb, and 8 (29.6%) with grade C or above spirometry readings straddled 70% for FEV_1_/FVC ratio. In those with asthma, the inter-individual variation in log (FeNO) accounted for 85.6 [77.7-91.1]% of total variance observed, 7.4 [4.5-12.5] % variance was observed within day and 6.6 [3.1-12.6] % between-day; this is similar for FEV_1_ % predicted (inter-individual: 78.5 [55.9-88.1]%, between-day: 10.9 [4.3, 23.0]% and within-day 10.7 [3.8-27.4]%). Whilst FEV_1_ % predicted and FEV_1_/FVC measured between 2PM and 8PM were higher than at other time points, FeNO was higher between 8 AM and 2 PM (Figures E17-E22).

**Figure 2.**
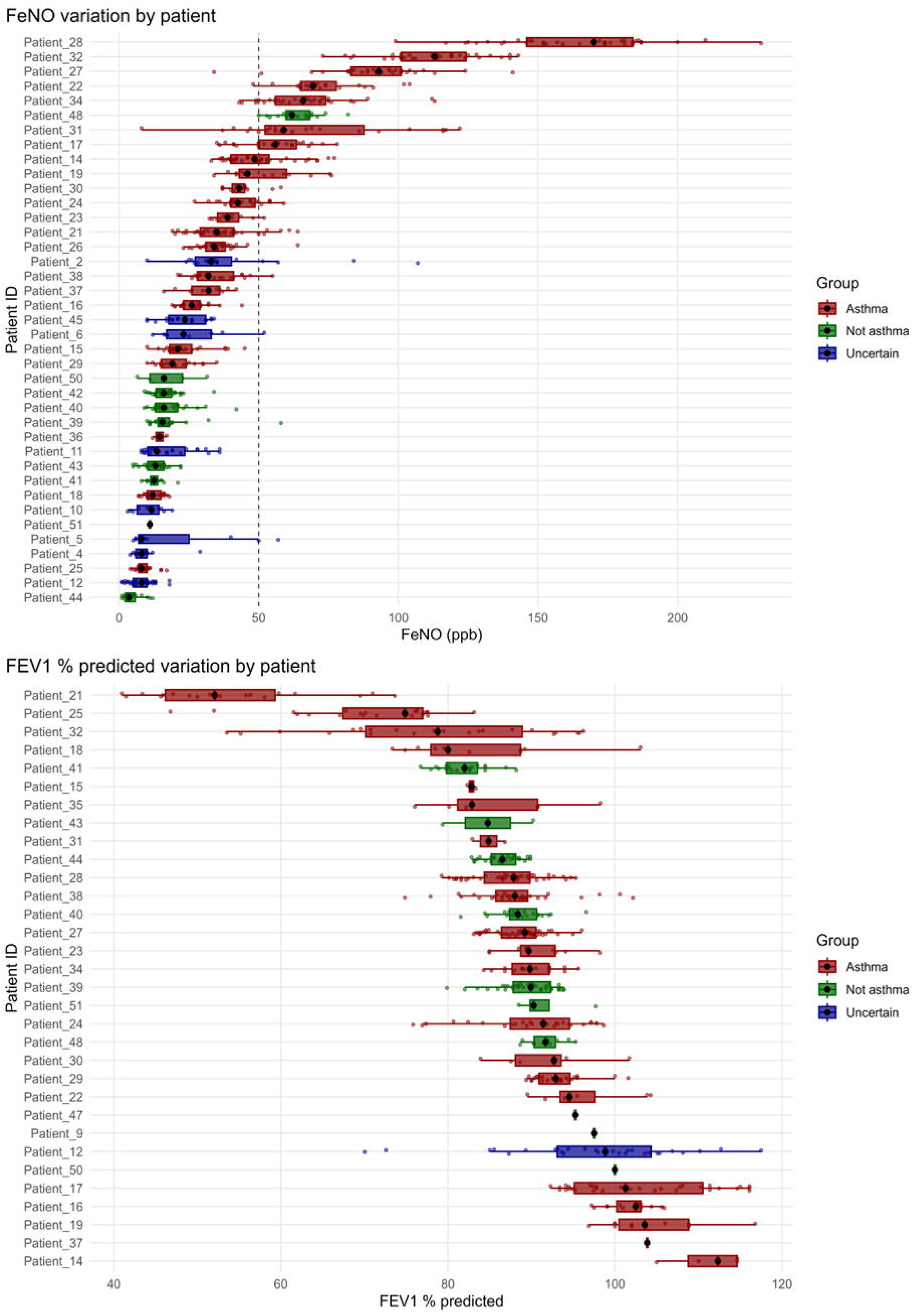
Intra- and inter- individual variability of home FeNO and FEV_1_ % predicted.

Increased symptoms weakly correlated with reduced FEV_1_ % predicted (r=-0.22 [95%CI: - 0.32, 0.11], p<0.001), FVC % predicted (-0.121 [-0.228, -0.012], p=0.030) and FEV_1_/FVC (-0.17 [-0.27, -0.01], p=0.003), but not with FeNO (p=0.552). FeNO was correlated with FEV_1_ % predicted (0.14 [0.04, 0.23], p=0.009) and FVC % predicted (0.114 [0.013, 0.213], p=0.027).

Low symptom scores at time of testing (VAS <4) were associated with higher FEV_1_% predicted (β = 5.39, 95% CI [3.20, 7.57]; p < 0.001) and FVC% predicted (5.06 [2.91, 7.17], p< 0.001) compared to tests done during high symptom period (VAS ≥4).

#### Putative diagnostic parameters

Twenty-eight participants with a confirmed diagnostic outcome completed more than one home spirometry measurement, and 32 participants with a confirmed diagnostic outcome completed home FeNO testing; these participants were included in the analysis to estimate the potential diagnostic utility of the putative home-testing parameters.

The maximum fluctuation in observed home FEV_1_% predicted (FEV_1_%Pred_max-min_) was higher in those with asthma versus those without (p=0.016). The variations in FEV₁ % predicted was evaluated as it adjusts for individuals’ demographics and is a recommended parameter for assessing bronchodilator reversibility by the European Respiratory Society and American Thoracic Society technical standard (2022) ^22^. In contrast, variability metrics based on day-to-day PEF fluctuation showed poorer diagnostic performance (Table 2, Figure E23). Using higher quality spirometry measurements (grade B above, or Grade A only) did not show better discriminative abilities (Table E4).

**Table 2.** Diagnostic efficiencies of routine tests and home-based testing metrics in participants who had confirmed diagnostic outcome (asthma and not asthma, n=38).

| Potential parameters<sup>§</sup> | Number (n) | AUC<sub>ROC</sub> | Threshold for specificity to be >90% for home tests; or established threshold for routine tests | Sensitivity | Specificity |
| --- | --- | --- | --- | --- | --- |
| **Home-based testing** |  |  |  |  |  |
| **Home spirometry parameters** |  |  |  |  |  |
| FEV<sub>1</sub>%Pred<sub>max-min</sub> (%) | 28 | 0.78 (0.62-0.95) | ≥16% | 52.4 (28.6-71.4)% | 100 (100-100)% |
| PEF<sub>max-min</sub>%, (%) | 28 | 0.61(0.38-0.84) | ≥50% | 19.1 (4.7-38.1)% | 100 (100-100)% |
| **Home FeNO parameters** |  |  |  |  |  |
| FeNO<sub>median</sub> (ppb) | 32 | 0.82 (0.65-0.99) | ≥25 | 77.3 (59.1, 95.5)% | 90.0 (70.0- 100.0)% |
| FeNO<sub>median_8AM-2PM</sub> (ppb) | 32 | 0.81 (0.63-0.99) | ≥21 | 81.8 (63.6, 95.5)% | 90.0 (70.0- 100.0)% |
| FeNO<sub>maximum</sub> (ppb) | 32 | 0.80 (0.62, 0.97) | ≥59 | 54.5 (31.8-72.7)% | 90.0 (70.0- 100.0)% |
| FeNO<sub>maximum_8AM-2PM</sub> (ppb) | 32 | 0.82 (0.66-0.99) | ≥43 | 72.7 (54.6-90.9)% | 90.0 (70.0- 100.0)% |
| FeNO<sub>IQR_variability</sub> (ppb) | 32 | 0.76(0.58, 0.93) | ≥12 | 50.0 (27.3-68.2)% | 90.0 (70.0- 100.0)% |
| **Routine tests** |  |  |  |  |  |
| Bronchodilator reversibility<sup>∞</sup> (% change in FEV<sub>1</sub>) | 37 | 0.93 (0.85-1) | ≥12% | 32.0 (16.0-52.0)% | 100 (100-100)% |
| Bronchodilator reversibility<sup>∞</sup> (% change in FEV<sub>1</sub> % predicted) | 37 | 0.93 (0.86-1) | ≥10% | 28.0 (12.0-48.0) | 100 (100-100)% |
| Blood eosinophil count<sup>∞</sup> (x10<sup>9</sup> cells/L) | 35 | 0.76 (0.60-0.92) | ≥0.4 | 45.5 (22.7-68.2)% | 100 (100-100)% |
| FeNO (NIOX)<sup>∞</sup> (ppb) | 38 | 0.80 (0.65-0.94) | ≥50 | 44.0 (24.0-64.0)% | 92.3 (76.9-100)% |
| FeNO (NOBreath) (ppb)<sup>Ω</sup> | 32 | 0.77 (0.58-0.96) | ≥50 | 31.8 (13.6-50.0)% | 90.0 (70.0-100)% |
| Methacholine PD<sub>20</sub><sup>∞</sup> (mg) | 31 | 1.00 (1.00 - 1.00) | <0.2 | 71.4 (52.4-90.5)% | 100 (100-100)% |
| <sup>§</sup>Equations:<br>FEV<sub>1</sub>%Pred<sub>max-min</sub>: maximum FEV<sub>1</sub>% predicted – minimum FEV<sub>1</sub>% predicted;<br>PEF<sub>max-min</sub>%, (maximum PEF – minimum PEF)/ minimum PEFx100;<br>FeNO<sub>median</sub>: Median of all home FeNO measurements;<br>FeNO<sub>median_8AM-2PM</sub>: Median of all home FeNO measurements performed between 8AM and 2PM;<br>FeNO<sub>maximum</sub>: Maximum of all home FeNO measurements;<br>FeNO<sub>maximum_8AM-2PM</sub>: Maximum of all home FeNO measurements performed between 8AM and 2PM;<br>FeNO<sub>IQR_variability</sub>: Interquartile range of all home FeNO measurements;<br><sup>β</sup>Upper limit of normal for local laboratory reference range; <sup>∞</sup> Diagnostic outcomes were taken into account of NIOX FeNO measurements, BDR, BEC and BCT results; <sup>Ω</sup> In-clinic NOBreath measurements were blinded to the Expert Panel when confirming diagnostic outcomes.<br>PD20: provoking dose of methacholine in inducing 20% fall in FEV<sub>1</sub>. |  |  |  |  |  |

All home FeNO-derived parameters were higher in individuals with asthma compared with those without (p<0.05, Figure E24) and had high AUC and sensitivity whilst maintaining specificity of >90% (Table 2, Figures E25).

#### Comparison with routine tests

A median (IQR) of 9.0 (6.0-11.8) days of diurnal home PEF (PEFv) data were obtained from 34 (66.7%) participants, irrespective of spirometry quality Grade. Of the 34 participants, 28 had confirmed diagnostic outcomes. Only four of these completed ≥14 days of twice-daily peak flow readings; all four had asthma, but none demonstrated ≥20% mean diurnal PEF variability. All clinic-based routine tests demonstrated high specificity (≥90%) using established cut off values (Table 2).

Clinic-based FeNO showed strong correlation with home FeNO_median_ (r=0.95 [0.90–0.98], p < 0.001) but only moderate agreement with 95% limits of agreement of -29.7 to +42.2 ppb (Figures E26). Lowering the diagnostic cut-off for clinic-based FeNO improved sensitivity but reduced specificity (69.2-84.6%, Table E5), depending on the threshold applied.

#### Integration into diagnostic pathway

Mapping the home-based diagnostic strategy onto the current BTS/NICE/SIGN 2024 adult pathway^16^ reduced the need for BCT by 65% (Figure 3), equivalent to offering home testing to 1.5 (95% CI [1.3–2.2]) patients to avoid one bronchial challenge test. Among 37 participants who had a confirmed diagnostic outcome who completed all BTS/NICE/SIGN 2024 pathway tests, the integrated home testing pathway achieved 94.6% accuracy (35/37) compared with 89.2% (33/37) for the current BTS/NICE/SIGN 2024 pathway, whilst leading to a reduction of 78.3% (18/23) who would have required BCT (Figure E27). Early economic scoping using NICE unit costs^26^ indicated that home testing would be cost-neutral on diagnostic costs alone if it reduced BCT by 25% (Table E6).

**Figure 3.**
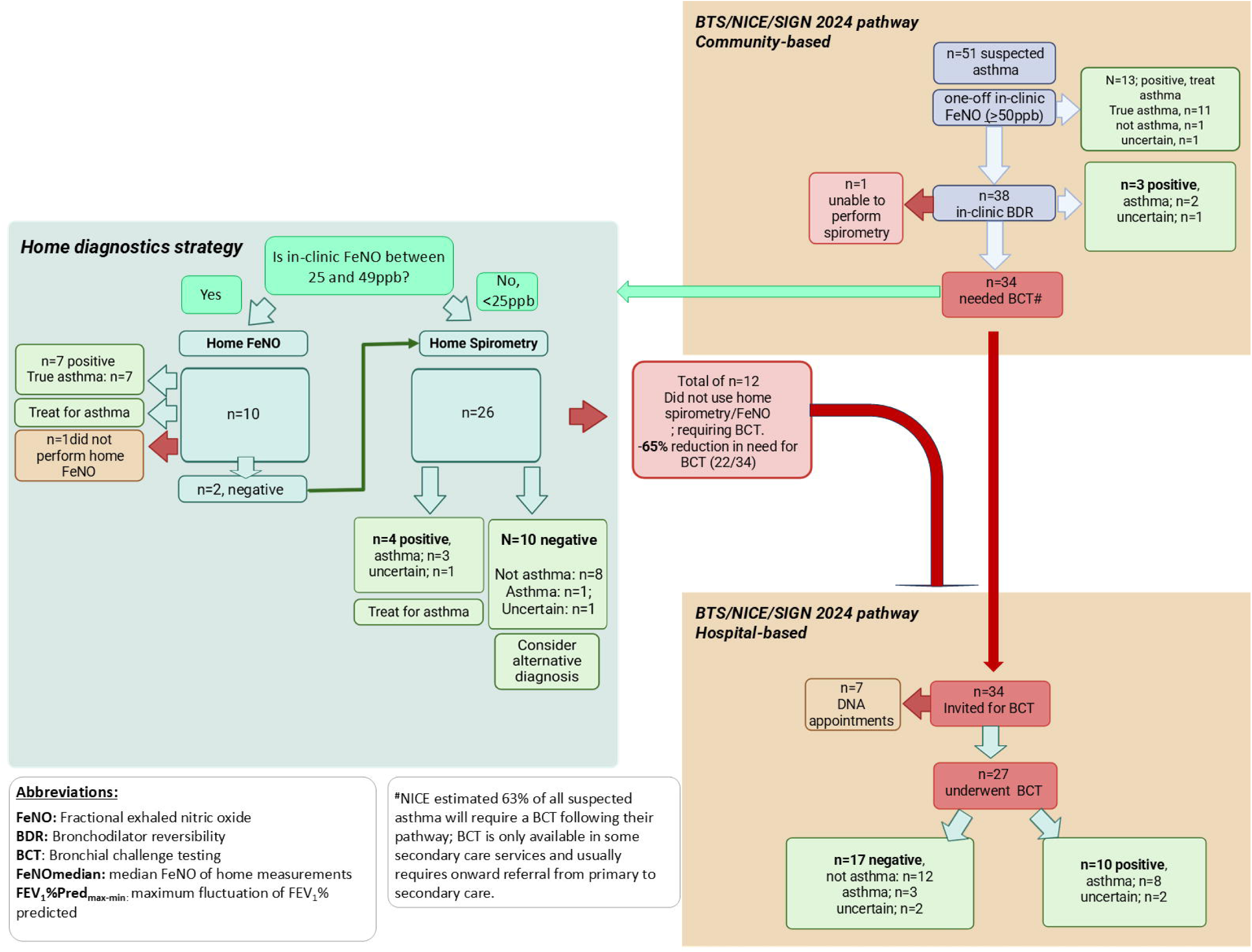
An example diagnostic strategy integrating home diagnostics into the BTS/NICE/SIGN 2024 adult asthma guideline pathway to reduce the need for bronchial challenge testing (BCT). This model begins with in-clinic FeNO and bronchodilator reversibility (BDR) testing (the first two steps of the BTS/NICE/SIGN 2024 pathway). For patients with negative in-clinic test results who would otherwise proceed to referral to secondary care for a BCT, home diagnostics are issued prior to discharge, guided by test results and technique quality. In patients with intermediate in-clinic FeNO (25-491ppb), additional longitudinal home FeNO monitoring may better capture overall airway inflammation than a single point measurement and therefore offers additional insight. In contrast, for those with low in-clinic FeNO (<251ppb), home spirometry may be a more appropriate option. Training for techniques in performing FeNO and spirometry could be delivered at the same time as patients are performing clinic-based tests, avoiding additional appointments or training time. Devices are returned after 1-2 weeks, with results reviewed and actioned remotely by healthcare professionals. This potential community-based one-stop-shop approach substantially reduces the number of patients who need referral to secondary care for BCT; in this cohort, 65% BCT could be avoided.

## DISCUSSION

Home-based spirometry and FeNO testing are feasible at the diagnostic setting. The putative metrics in home testing generated better diagnostic yield than home PEFv; integrating these tests into the current BTS/NICE/SIGN 2024 diagnostic pathway could reduce the need for BCT, with the potential to provide value for money.

Short term longitudinal diagnostic testing is established in other conditions, such as hypertension, where ambulatory blood pressure monitoring reduces misdiagnosis arising from reliance on single clinic-based measurements and is now embedded in routine practice.²⁷ Asthma biomarkers are similarly dynamic.¹⁰ Consistent with our previous work,^11,12,15^ we observed marked temporal variability in FEV_1_ and FeNO, despite no change in treatment. Variability in home FEV₁ may reflect bronchial hyperresponsiveness to everyday stimuli, bronchodilator effects following reliever use when symptomatic, circadian influences^28^, or fluctuations in airway inflammation^10^, underscoring the importance of longitudinal measurements for asthma diagnosis.

To date, diurnal PEF variability remains the only widely available, low-cost home-based diagnostic tool for asthma diagnosis^9,16^. However, PEFv measurements and interpretations remains problematic: adherence to twice-daily measurements, suboptimal test technique and challenges with accurate documentation of results using non-digital tools limits its clinical utility^29,30^. Furthermore, PEFv demonstrates poor diagnostic sensitivity (∼15%)^31^, reflecting its limited ability to detect airflow obstruction arising from the smaller airways^32^. Digital approaches using more sensitive biomarkers (such as FEV₁) may therefore improve both test adherence and diagnostic yield^34^.

The diagnostic utility of FeNO and the appropriate threshold for asthma diagnosis remain debated across major guidelines.⁷ The substantial overlap in FeNO values between individuals with and without asthma may partly reflect unaccounted temporal variability.³⁵ Repeated FeNO measurements during the diagnostic process may better represent an individual’s Type-2 inflammatory burden, improve diagnostic certainty and enable the use of a lower diagnostic threshold. The observed variability also suggests that repeated measurements may be required when establishing a minimally clinically important difference in FeNO-based treatment response.³⁶ Future studies should investigate whether home FeNO testing improves diagnostic accuracy, particularly among patients with indeterminate one-off clinic-based FeNO results.

It is important to note that data from the RADicA study contributed to the development of the BTS/NICE/SIGN 2024 diagnostic pathway.²⁶ The current study used the same design, enabling direct evaluation of how home-based testing could integrate into this pathway.

Although the current prospective data are independent of those shared with NICE, the estimated requirement for BCT was similar (65% vs 63% ^26^); this highlights a major implementation bottleneck, given that BCT is largely restricted to secondary care. Novel diagnostic technologies and strategies that safely reduce reliance on BCT would therefore be of clear value. However, establishing the viability of home-based testing at scale will require an understanding of barriers and facilitators to uptake among key stakeholders,^17,18^ alongside validation in their clinical utility and formal health-economic evaluation.

### Limitations

This study is limited by its small sample size, reflecting its feasibility design and resulting in wide confidence intervals. Nevertheless, feasibility was evaluated in a setting representative of real-world diagnostic practice, including a demographically diverse population, with most participants recruited from highly deprived areas.

In-clinic FeNO measurements were performed following avoidance of caffeine, whereas this was not required during home testing for practical and adherence considerations. However, it is important to highlight that evidence regarding the effect of caffeine ingestion on FeNO remains controversial^37^.

Neither the FeNO device nor the home spirometer was designed specifically for diagnostic use at home. Accordingly, this was not a device validation study but an evaluation of participant ability to perform home testing and the potential clinical utility of these measurements. Nevertheless, our findings highlight new opportunities for device optimisation for diagnostic use. It is also important to note that the four-times-daily testing schedule was used for feasibility evaluation and is unlikely to be the strategy in future studies; our findings suggest that targeting specific time-of-day windows and symptom periods could substantially reduce testing burden while maintaining diagnostic accuracy.

## CONCLUSION

Home spirometry and FeNO testing were feasible and showed early potential to improve asthma diagnosis. An adequately powered diagnostic accuracy study is urgently needed to validate these approaches. If validated, uptake and adherence in routine care could be enhanced through improved device design, clearer outputs and appropriate implementation support, followed by evaluation of clinical and cost-effectiveness.

## Supporting information

OLS

## Data Availability

The authors will consider all reasonable requests for deidentified data, following approval by the study sponsors. Proposals should be directed to. To gain access, data requestors will need to sign a data access agreement.

## Acknowledgement

We would like to thank the RADicA study participants for their time and commitment to the study and the RADicA study team for help with data collection.

RW, MB, AS, SJF, HD and CSM are supported by the NIHR Manchester BRC. RW is supported by NIHR Clinical Lectureship (CL-2023-06-003). HJD is supported by MRC-CSF-MR/V029460/1

