## Supplementary material for "Can home spirometry and FeNO testing improve asthma diagnosis? – a feasibility study": OLS

**Online supplementary materials**

***Table E1.*** *Baseline characteristics of recruited patients.*

|  | **Asthma**  **(n=25)** | **Not asthma**  **(n=13)** | **Uncertain diagnosis**  **(n=13)** |
| --- | --- | --- | --- |
| Male, n (%) | 11 (44%) | 7 (50.0%) | 3 (25%) |
| Age, yrs, median (IQR) | 30 (24-39) | 22 (20-32) | 37 (28-39) |
| Ethnicity, white, n(%) | 15 (62.5%) | 7(53.8%) | 6 (46.2%) |
| Digital confidence (confident), n (%)^Ω^ | 17 (77.3%) | 11 (84.6%) | 5/10 (50%) |
| Deprivation index (IMD decile≤3), n (%) | 15 (60%) | 8 (61.5%) | 9 (69.2%) |
| Income Decile (<3), n (%) | 15 (60.0%) | 7 (53.8%) | 8 (61.5%) |
| Education and skills decile (<3), n(%) | 11 (44%) | 5 (33.3%) | 6 (54.5%) |
| Health and disability decile(<3), n(%) | 21(84%) | 12 (92.3%) | 11 (84.6%) |
| Barriers to Housing and services Decile (<3), n(%) | 9 (36.0%) | 6 (46.2%) | 7 (53.8%) |
| Living environment decile (<3), n(%) | 9 (36.0%) | 8 (61.5%) | 4 (30.7%) |
| Body mass index , kg/m^2^, median (IQR) | 27.1 (24.4-33.6) | 25.7 (23.8, 28.7) | 35.6 (34.2, 39.1) |
| ACQ-5 points, median (IQR) | 1.4 (0.6-2.4) | 1.4 (1-2.2) | 1.8 (1.0-2.8) |
| Self-reported diurnal variation in symptoms*, n (%) | 14/22 (63.6%) | 9/ 14 (64.3%) | 7/ 9 (77.8%) |
| Self-reported day-to-day variation in symptoms**, n (%) | 7/24 (30.4%) | 6/12 (50%) | 7 (53.8%) |
| FEV_1_ % predicted, %, median (IQR) | 92.0 (87.5 – 99.4) | 101.0 (94.9-107) | 97.3 (92.4, 106) |
| FEV_1_/FVC, % median (IQR) | 74.7 (70.2-78.5) | 82.3 (79.8, 86.9) | 79.4 (77.5, 83.0) |
| Bronchodilator reversibility, %, median (IQR) | 8 (5-11) | 2 (0-4) | 2.5 (-1.3, 5)  N=12 |
| Positive methacholine^#^, n (%) | 15 (71.4%)  N=21 | 0  N=10 | 1 (25.0%)  N=4 |
| Positive mannitol challenge^$^, n (%) | 6 (60%),  N=10 | 1 (14.3%)  N=6 | 0  N=1 |
| At least one BCT positive*^∞^*, n (%) | 17 (73.9%)  N=23 | 0  N=13 | 2 (50%)  N=4 |
| FeNO, ppb, median (IQR) | 43 (25-69) | 19 (17-22) | 18(13-22) |
| Blood eosinophil counts, x10^9^ cells/L, median (IQR). | 0.30 (0.16-0.48)  N=24 | 0.13 (0.06-0.25)  N=11 | 0.14 (0-0.21)  N=10 |
| SPT sensitised, n (%) | 22 (95.7%)  N=23 | 5 (38.5%) | 4 (36.4%)  N=11 |

**“Do you have variation of your symptom through the day? Yes/No”*

*** “Over the past week, do you have variations of symptoms from day to day? Yes/No”*

*# defined as PD_20_<0.2mg; $ defined as PD_15_<635mg; ∞either methacholine and/or mannitol challenge positive.*

^Ω^ *All participants were asked the time of device training: “How confident do you generally feel about using digital products and mobile phone apps?” with five ordinal response options of “confident”, “somewhat confident”, “neutral”, “somewhat not confident”, and “not confident”.*

***Table E2.*** *The agreement between Expert Panel (enhanced reference standard) and BTS/NICE/SIGN 2024 pathways.*

|  |  | **Reference standard: Expert panel** | | |
| --- | --- | --- | --- | --- |
| **BTS/NICE/SIGN 2024 adult diagnostic pathway** |  | **Not asthma** | **Asthma** | **Uncertain** |
|  | **Not asthma** | *10* | *2* | *2* |
|  | **Asthma** | *1* | *23* | *4* |
|  | **Uncertain** | *2* | *0* | *7* |

***Figure E1.*** Home testing diary book

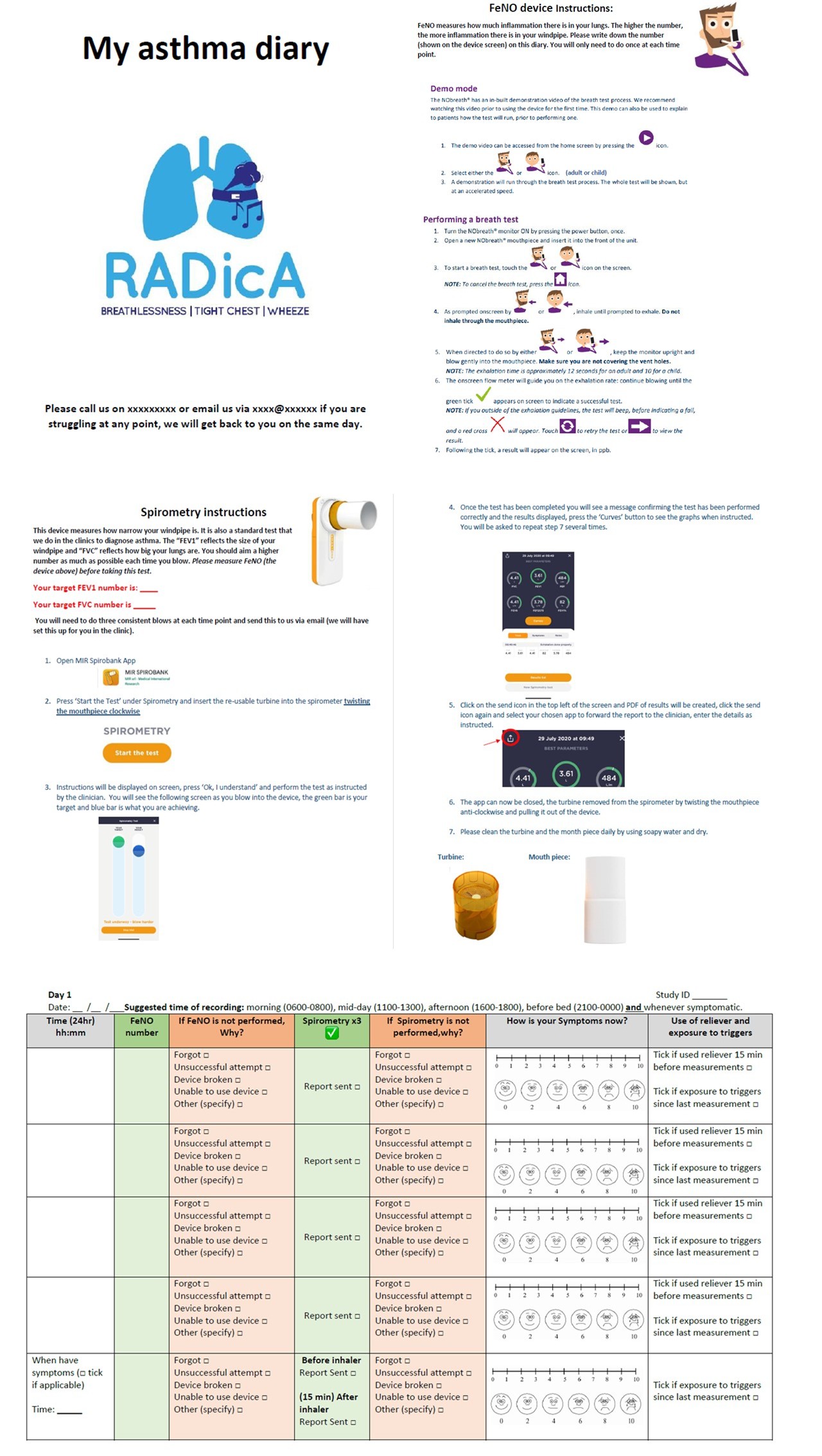

***Figure E2.*** Compliance rates for home spirometry and FeNO reporting during the testing period among participants with at least one completed assessment.

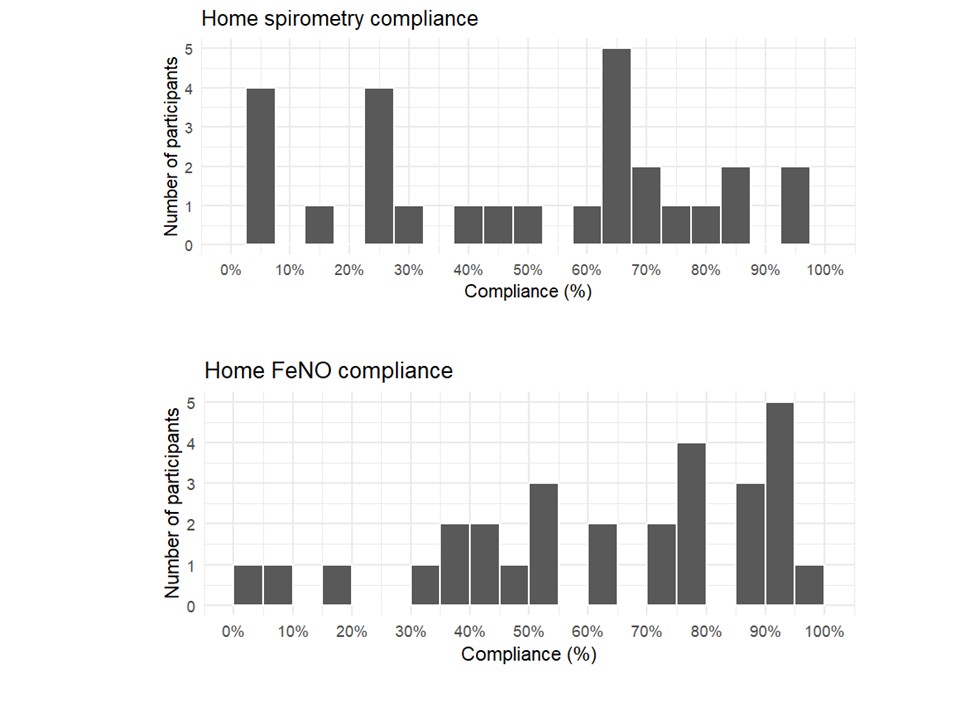

***Figure E3***. Quality of home spirometry (Grade A-F based on FEV_1_)

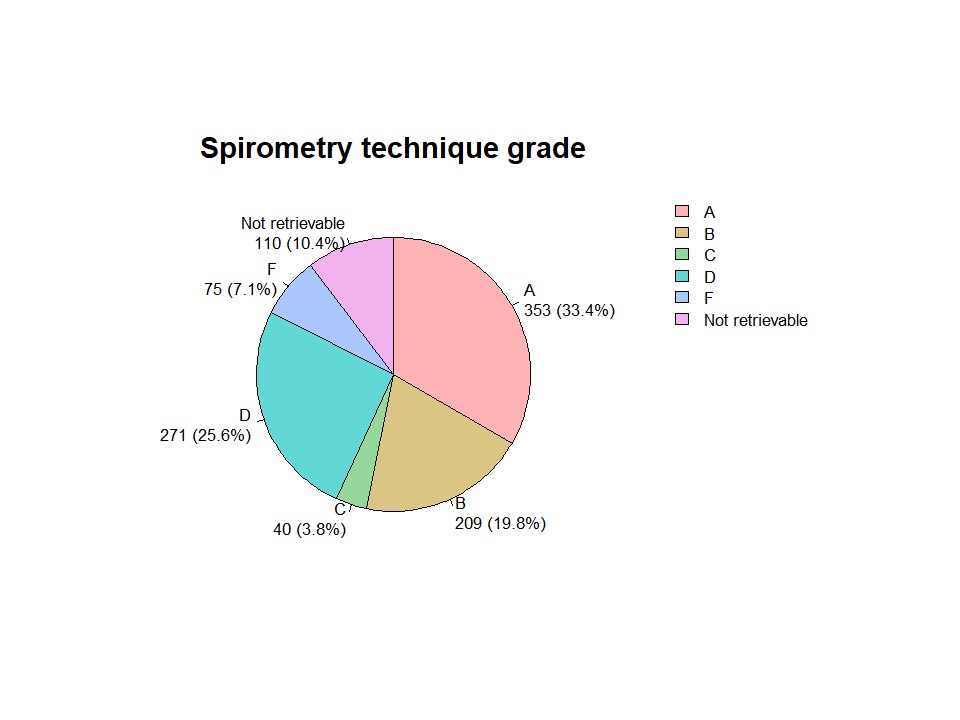

***Figure E4***. Home spirometry acceptability survey results
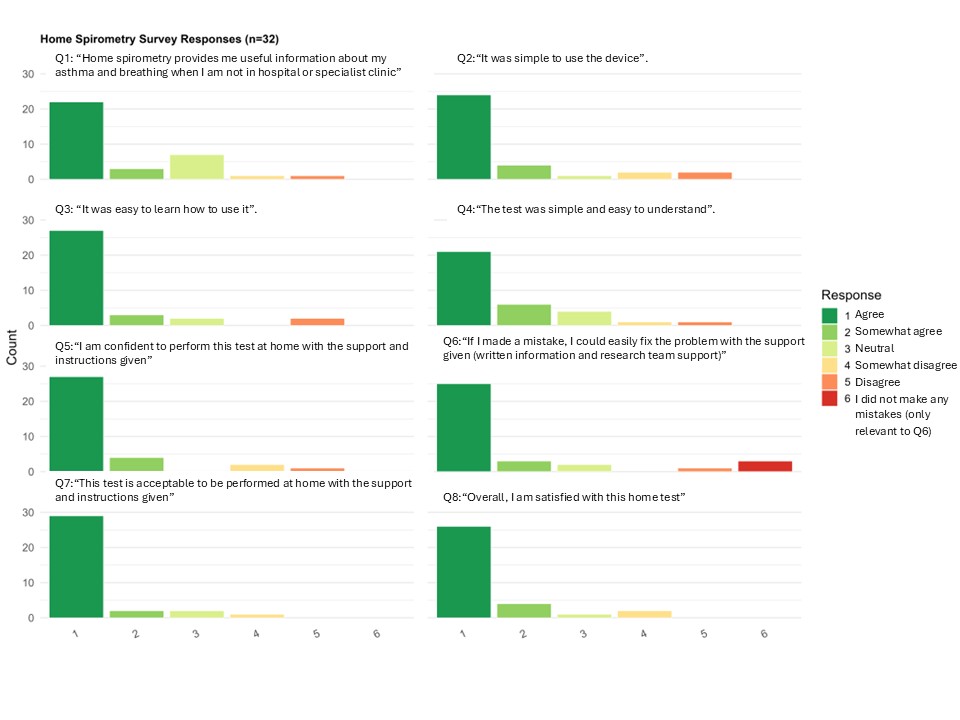

***Figure E5***. Home FeNO acceptability survey results

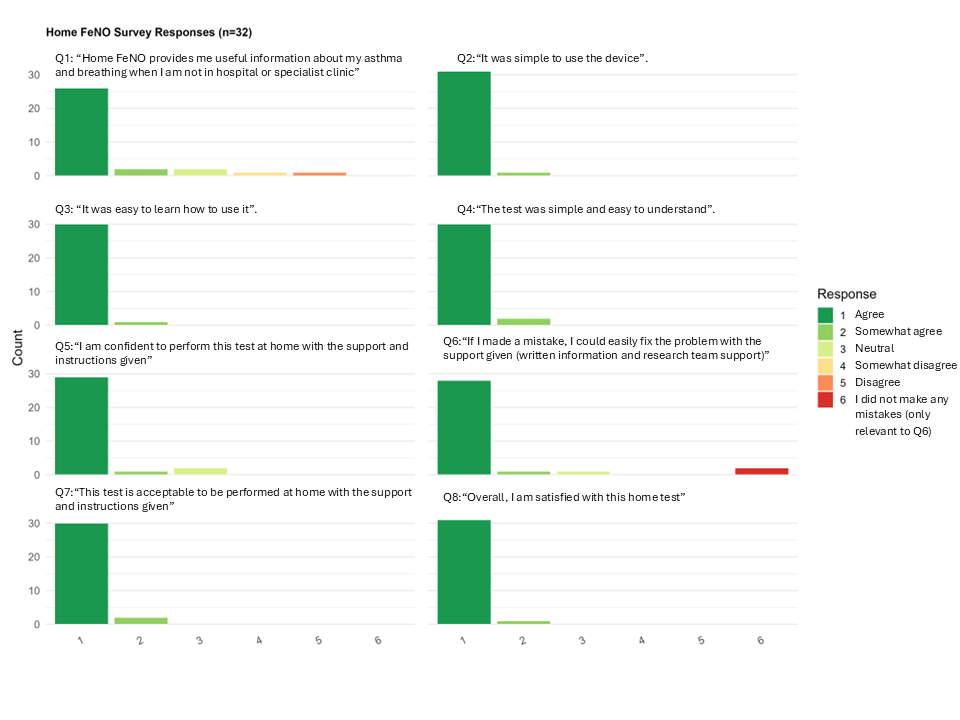

**Figure E6.** Daily EQ-5D-5L diary completion during home testing period declined from 62.7% (32/51) on day 1 to 19.6% (10/51) on day 14, with a mean daily completion rate of 43.0% across the 14-day monitoring period (Figure E6A). Participants from the most deprived areas (IMD deciles <3) reported lower mean utility than those from less deprived areas throughout the monitoring period (Figure E6B), though wide confidence intervals reflect small subgroup sizes. Participant engagement was bimodal: approximately one-third completed no diary days, while those who engaged typically completed 11 or more of the 14 days. When the diary was attempted, all five dimensions were consistently completed with no selective item non-response. Median (IQR) EQ-5D-5L utility index (England value set)^1^ was 0.940 (0.831-1.000), with 43.6% of observations at full health. Pain/discomfort was the most frequently reported problem (48% reporting any level of problems), followed by anxiety/depression (38%) and usual activities (36%). Panel A. EQ-5D diary completion during home testing period (14 days); n=51 participants, all 5 dimensions required. Dashed line indicates 50% completion threshold. Panel B. Mean EQ-5D-5L utility index over 14 days stratified by Index of Multiple Deprivation (IMD) group. Shaded areas represent 95% confidence intervals. Sample sizes are shown per timepoint.

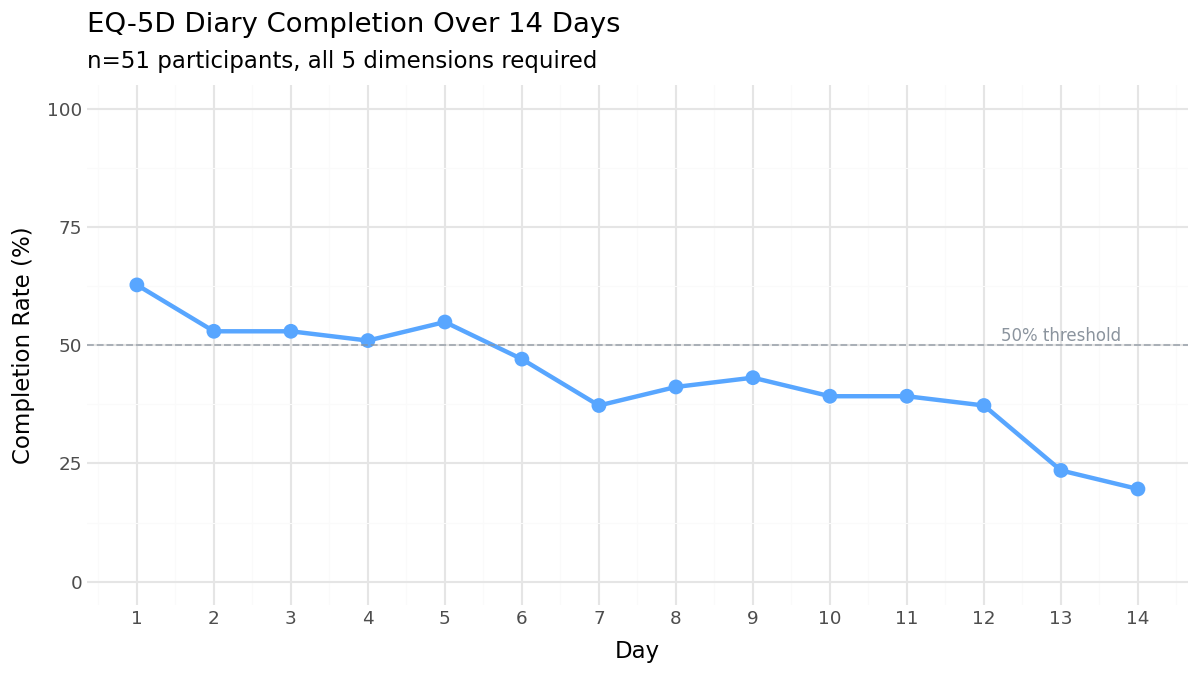

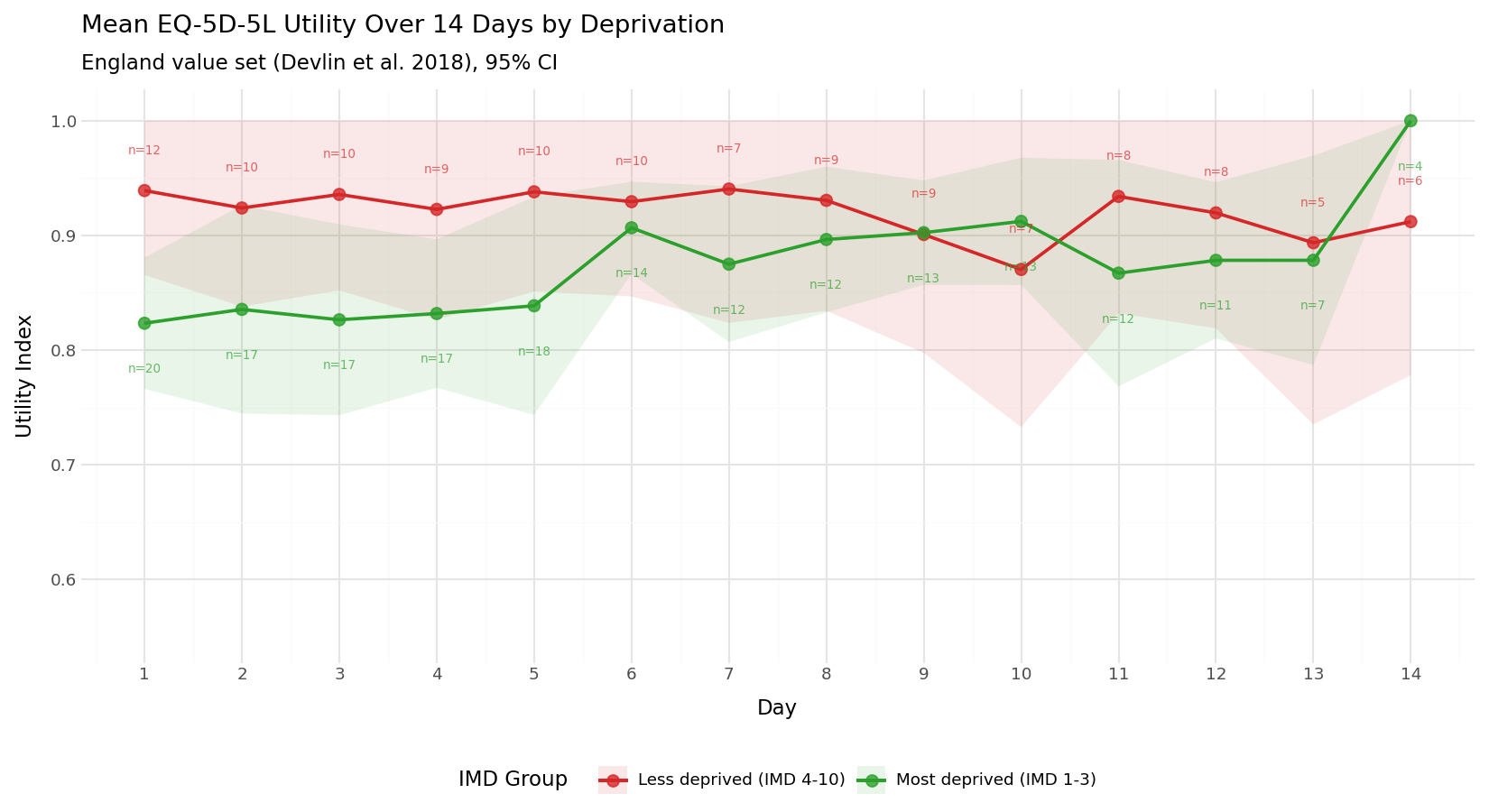

**Table E3.** EQ-5D-5L dimension response distributions**.** Pooled across 14 diary days, n=307 observations from 35 participants. England EQ-5D-5L value set. ^1^

| Dimension | No problems | Slight | Moderate | Severe | Extreme |
| --- | --- | --- | --- | --- | --- |
| Mobility | 81.6% | 12.3% | 5.8% | 0.3% | 0.0% |
| Self-Care | 88.0% | 11.4% | 0.6% | 0.0% | 0.0% |
| Usual Activities | 64.4% | 24.3% | 10.0% | 1.0% | 0.3% |
| Pain/Discomfort | 52.1% | 33.3% | 13.3% | 1.3% | 0.0% |
| Anxiety/Depression | 62.5% | 25.9% | 9.4% | 2.3% | 0.0% |

**Figure E7.** EQ-5D-5L dimension response distributions across the 14-day monitoring period.
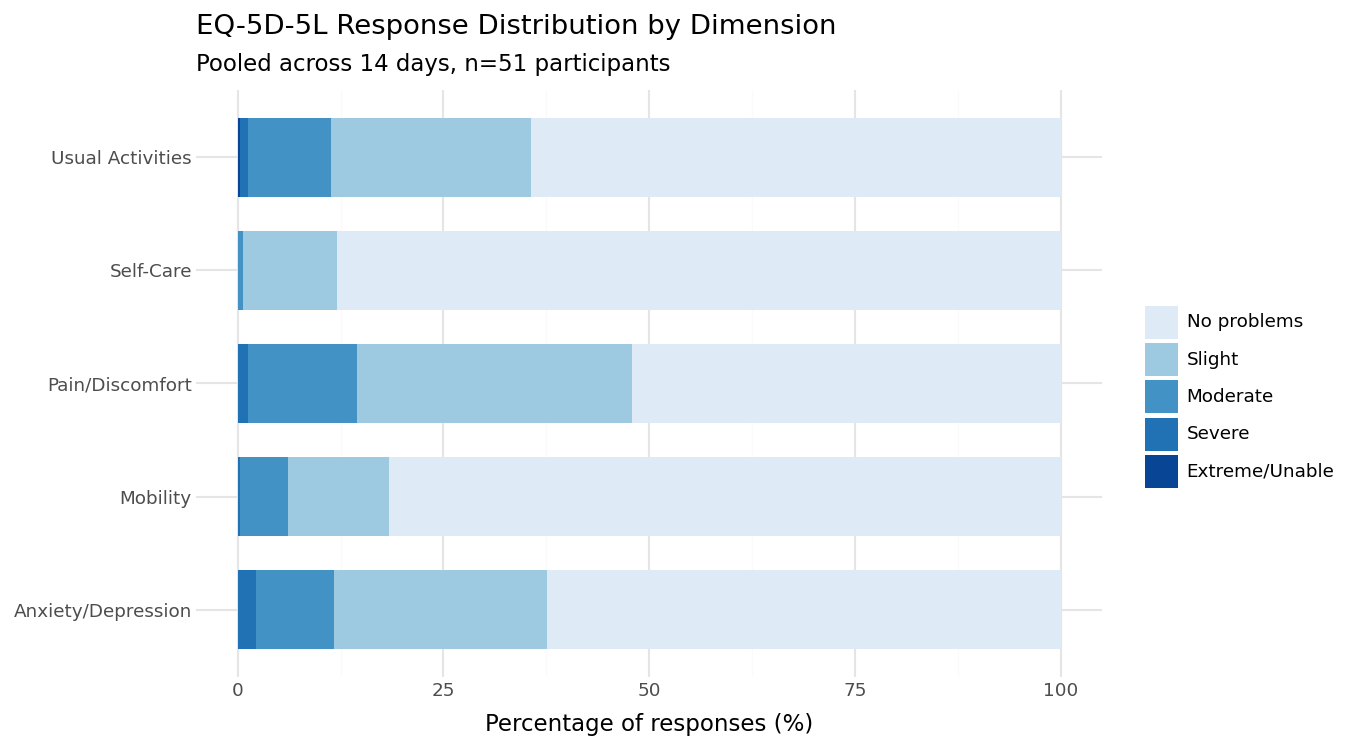

**Figure E8.** Feasibility of economic data collection across domains (n=51 participants). Bars show proportion of participants completing each measure. Dashed line indicates 80% completion threshold. Lower bars show unscheduled healthcare contact event rates rather than completion rates. 30% (11/37) of participants reported at least one unscheduled healthcare utilization due to respiratory symptoms within the previous 3 months, comprising unscheduled GP contacts (n=9), NHS 111 calls (n=3), and emergency department attendances (n=2). Overall, 71% of participants provided data sufficient for economic costing across all three domains (healthcare contacts, out-of-pocket costs, and productivity impact).

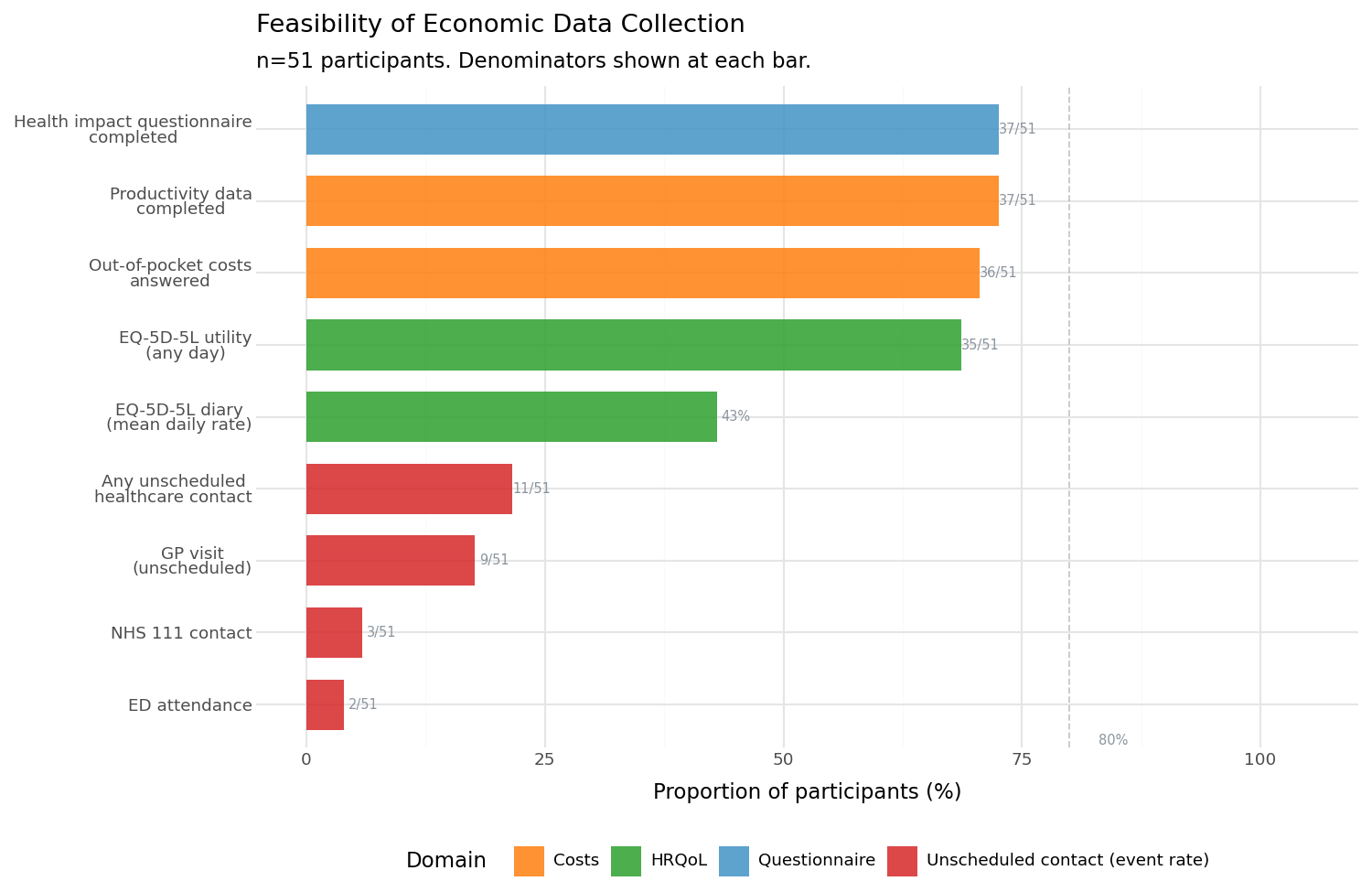

**Figure E9.** FeNO measurement by patient per day
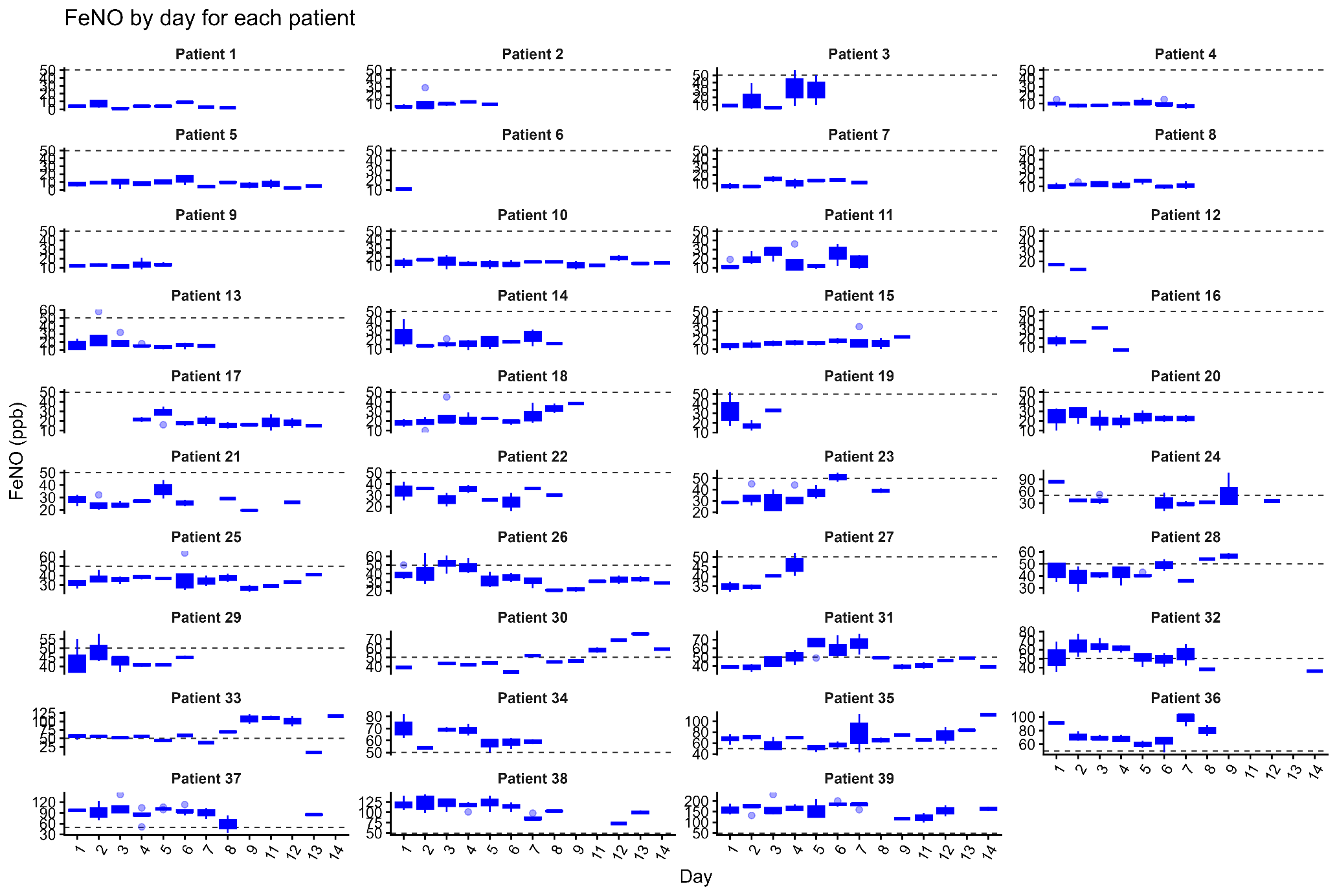

**Figure E10**. FEV_1_ % predicted variation by patient per day (only grade C or above spirometry included).

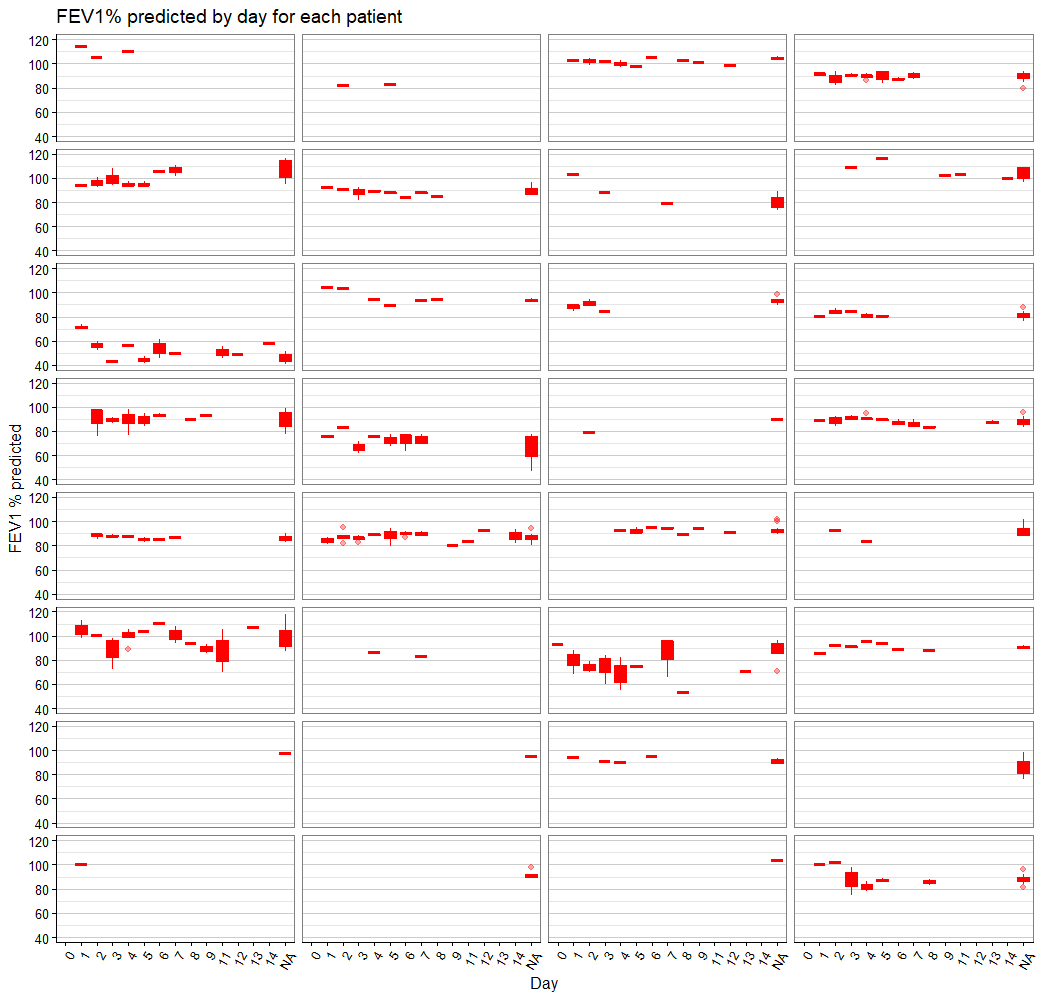

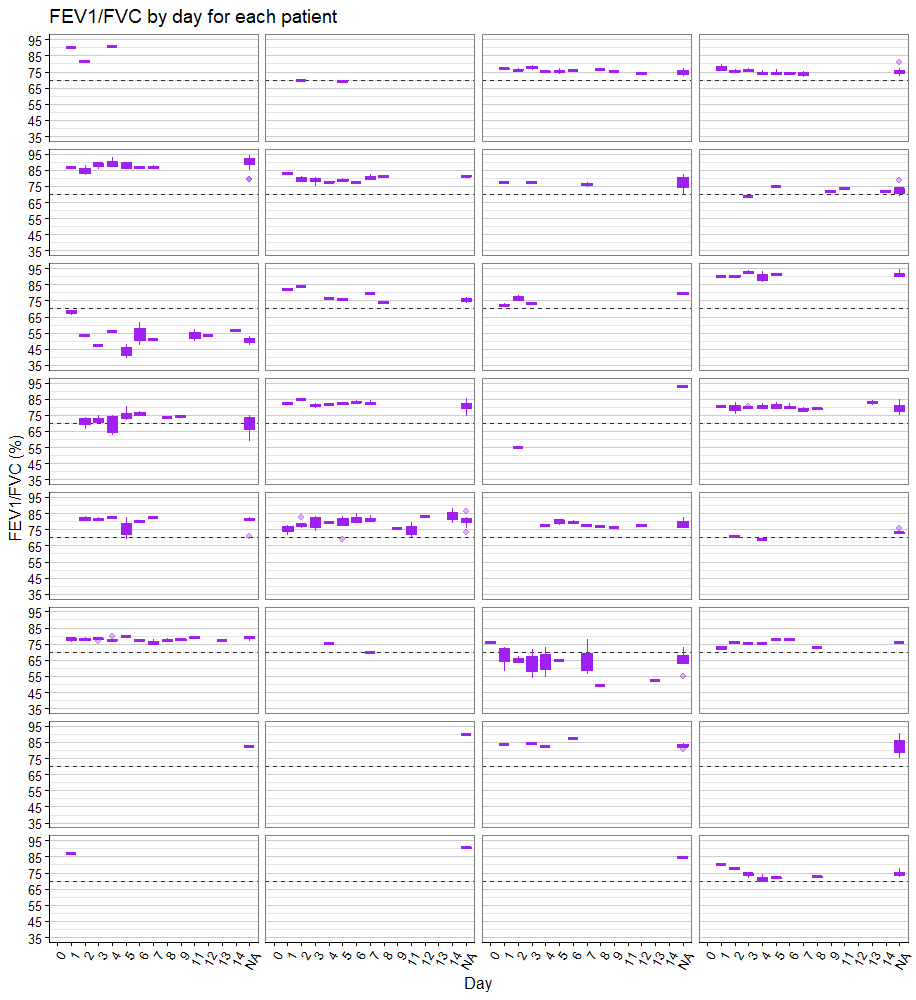
**Figure E11.** FEV_1_/FVC ratio variation by patient per day (only grade C or above spirometry included).

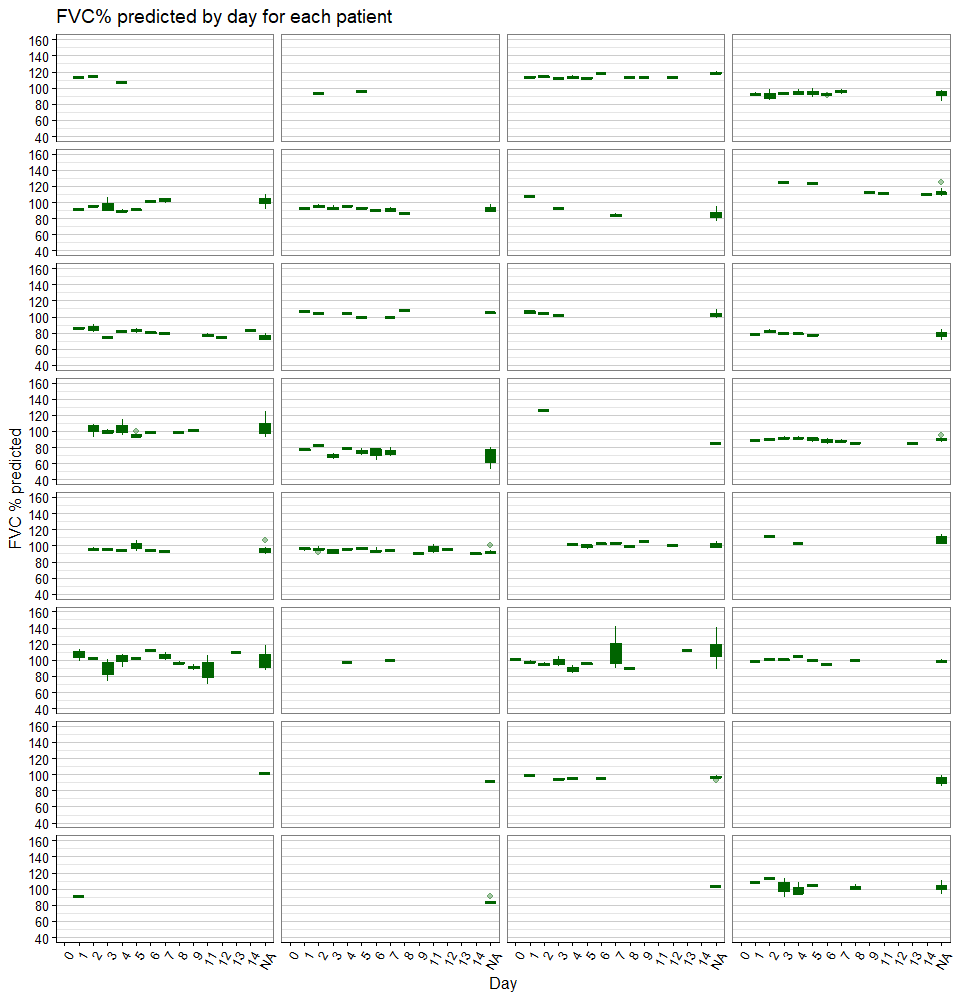
***Figure E12.*** FVC % predicted variation by patient per day (only grade C or above spirometry included).

***Figure E14.*** *Significant inter-and intra individual variation in home FEV1/FVC.*

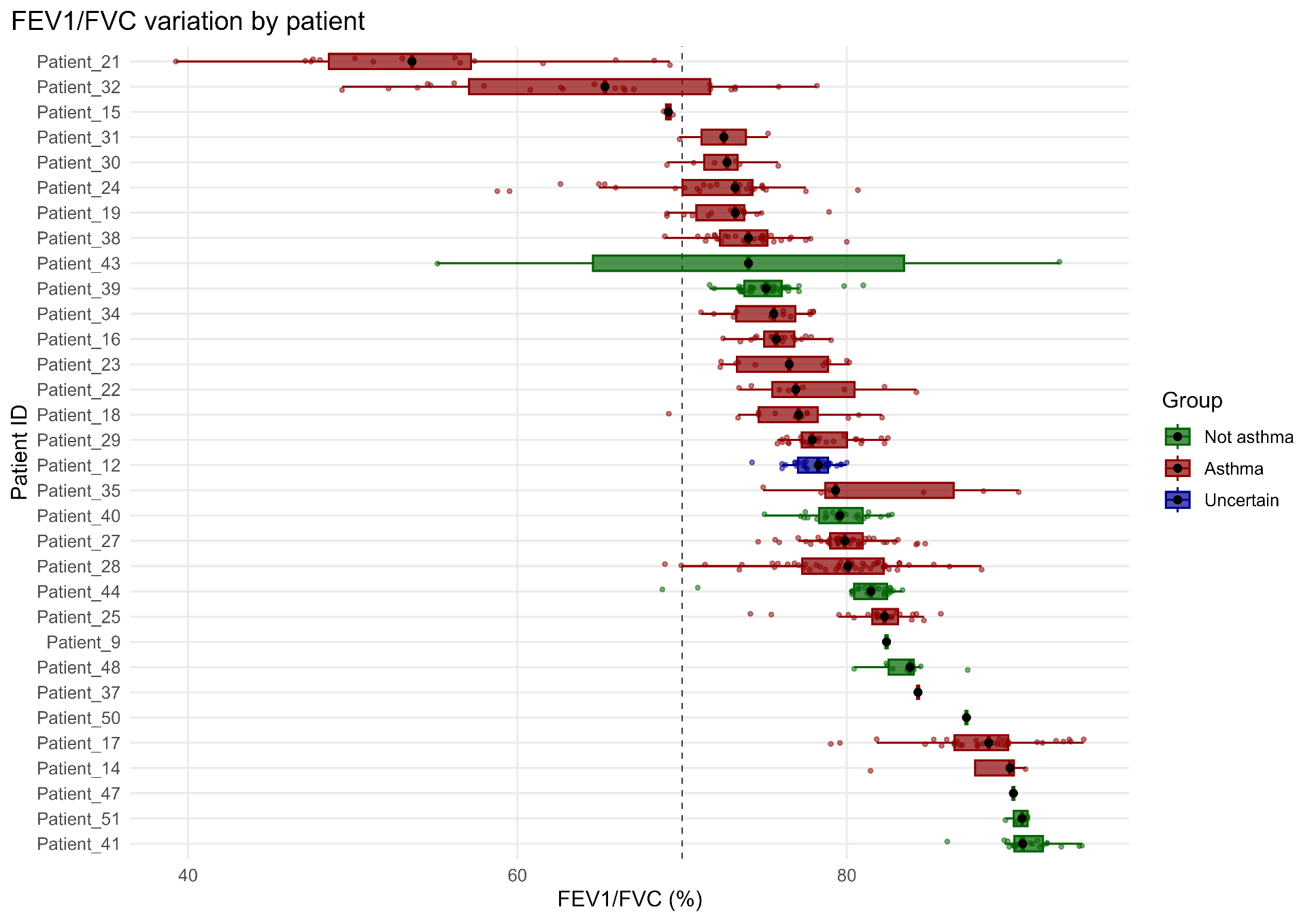

***Figure E15.*** *Significant inter-and intra individual variation in symptoms.*

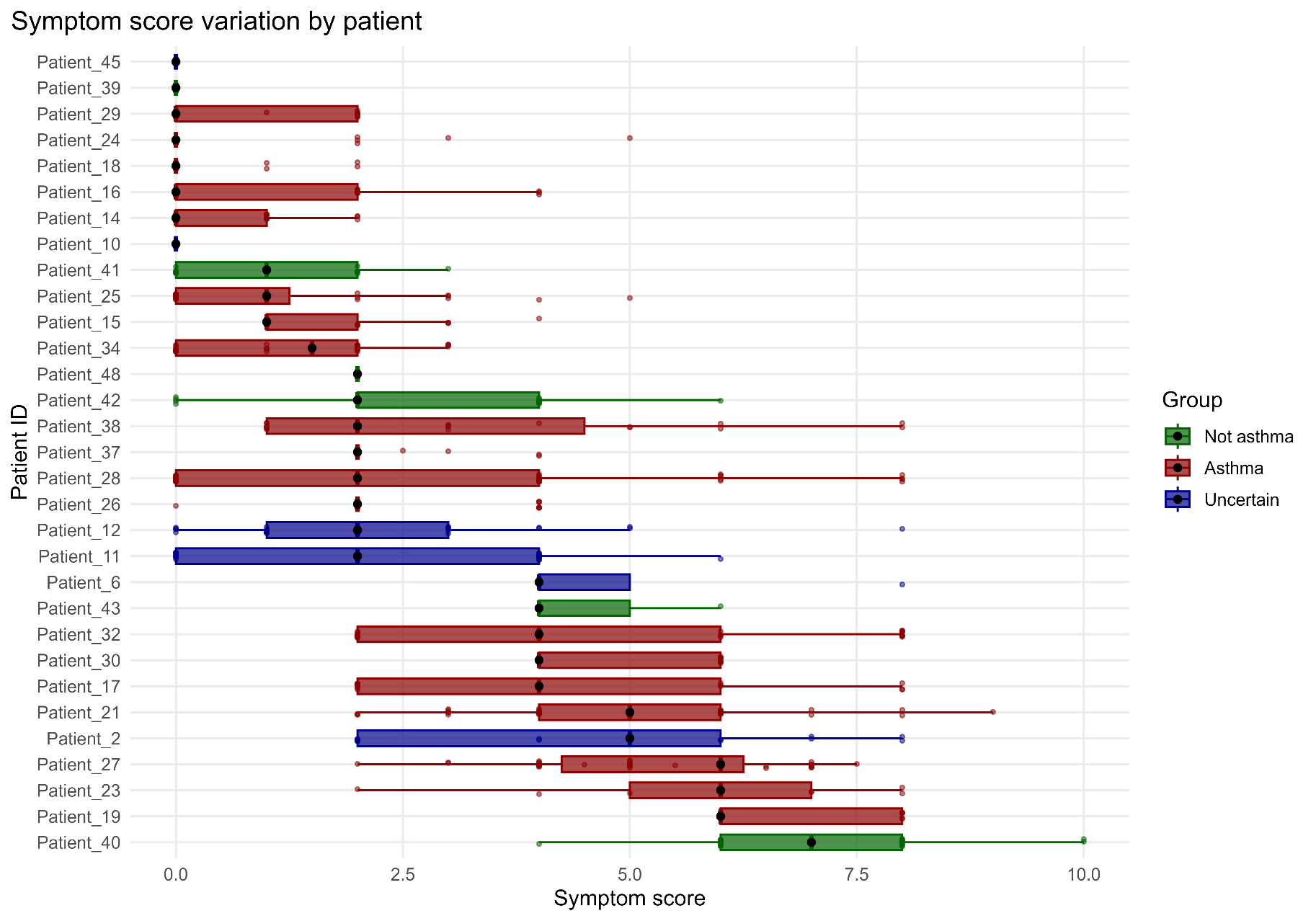

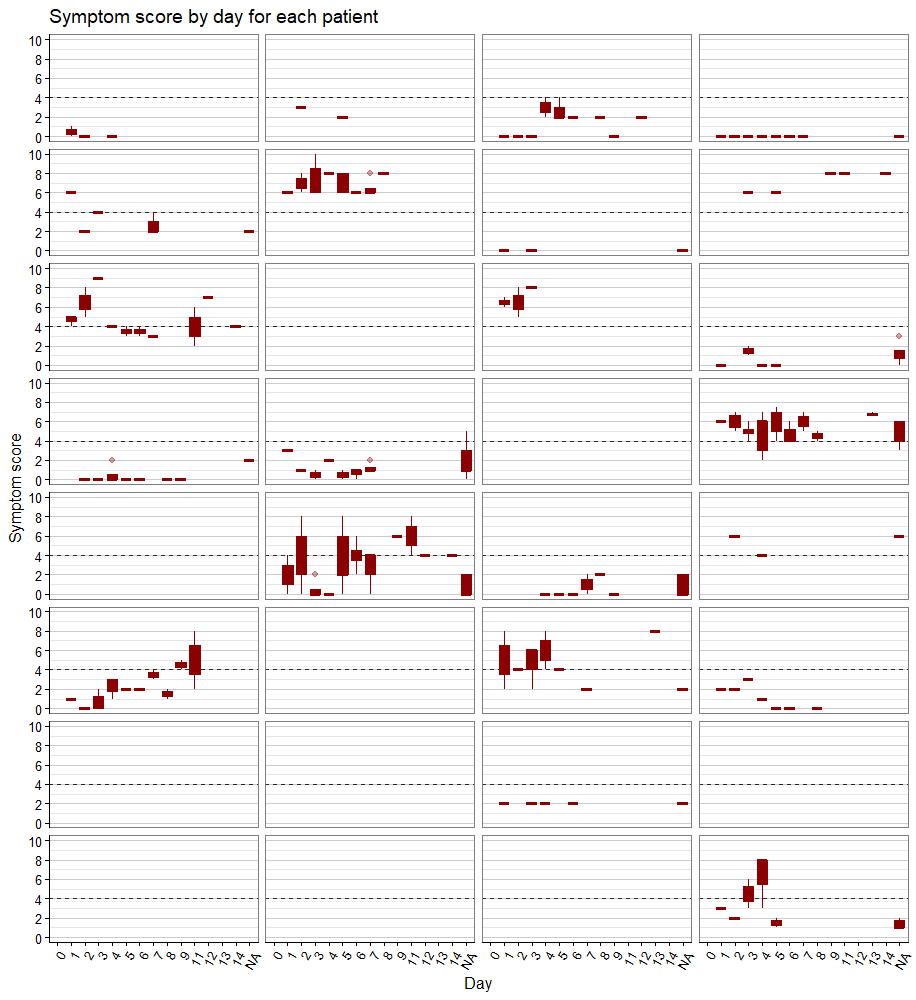
***Figure E16.*** VAS by day and participants.

**Time of day effect analysis:** Adjusted for repeated measures, within-day (diurnal) variations in home FeNO and spirometry measurements were explored by time of day by dividing the 24-hour period into four 6-hour time slots covering working hours (08:00–14:00 and 14:00–20:00) and resting hours (20:00–02:00 and 02:00–08:00).

***Figure E17.*** FeNO tend to be higher in the overall population, n=39 using mixed effect models.

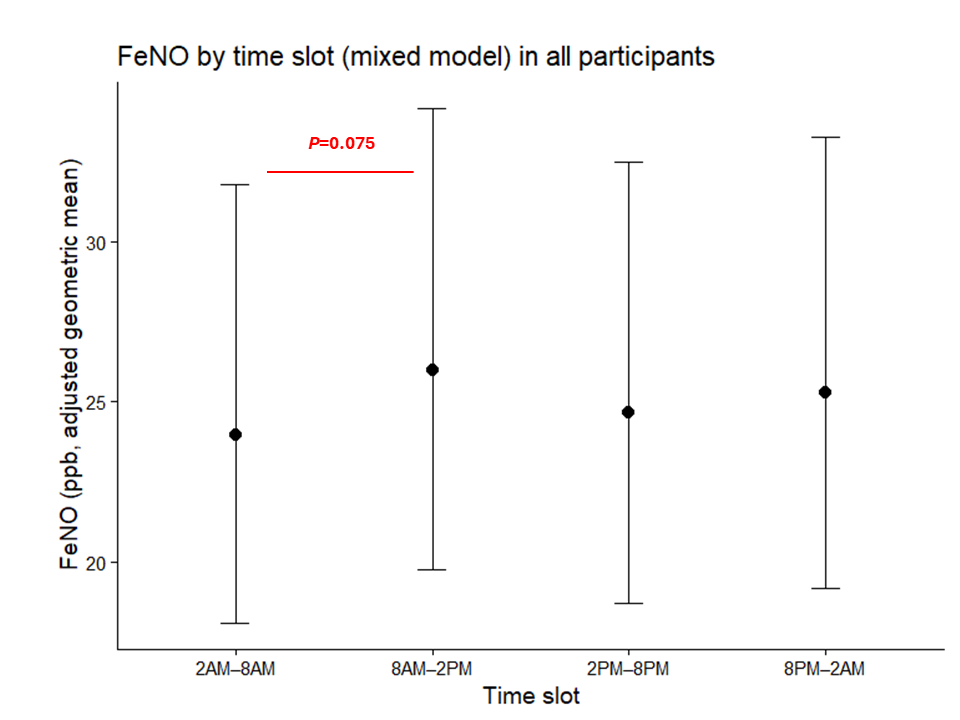

***Figure E18***. Association between time of day and log(FeNO) stratified by diagnostic group. A time-of-day trend was observed only in patients with asthma. Estimates were derived from a mixed-effects model adjusted for intra-individual repeated measures, with participant and day of measurement included as random effects (asthma: *p* = 0.047).

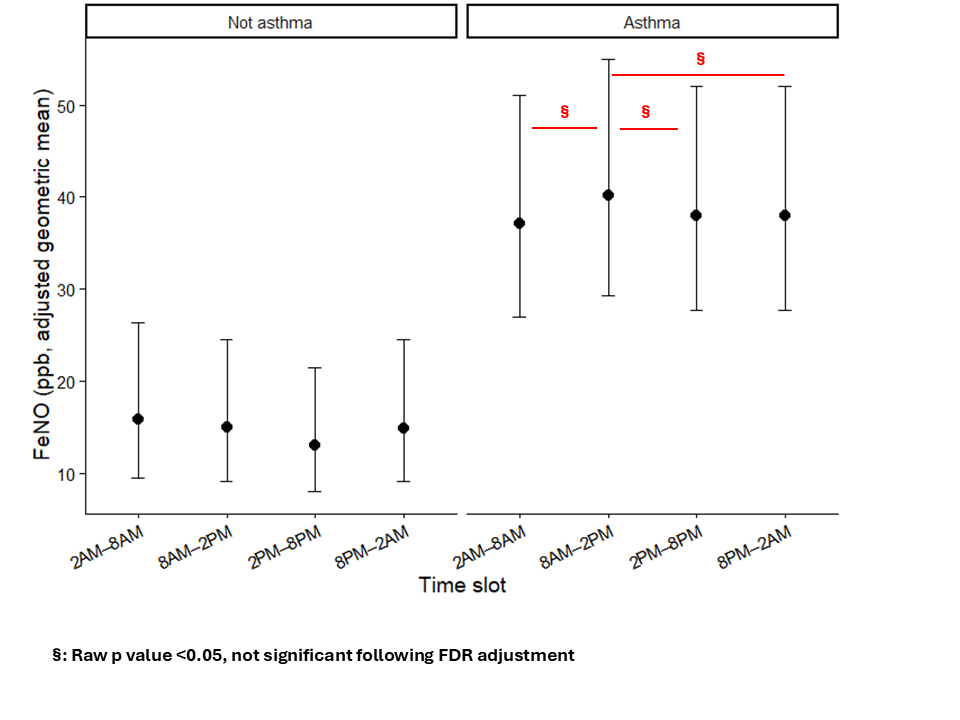

***Figure E19****.* FEV_1_ % predicted was higher in the overall population using mixed effect models, n=32.

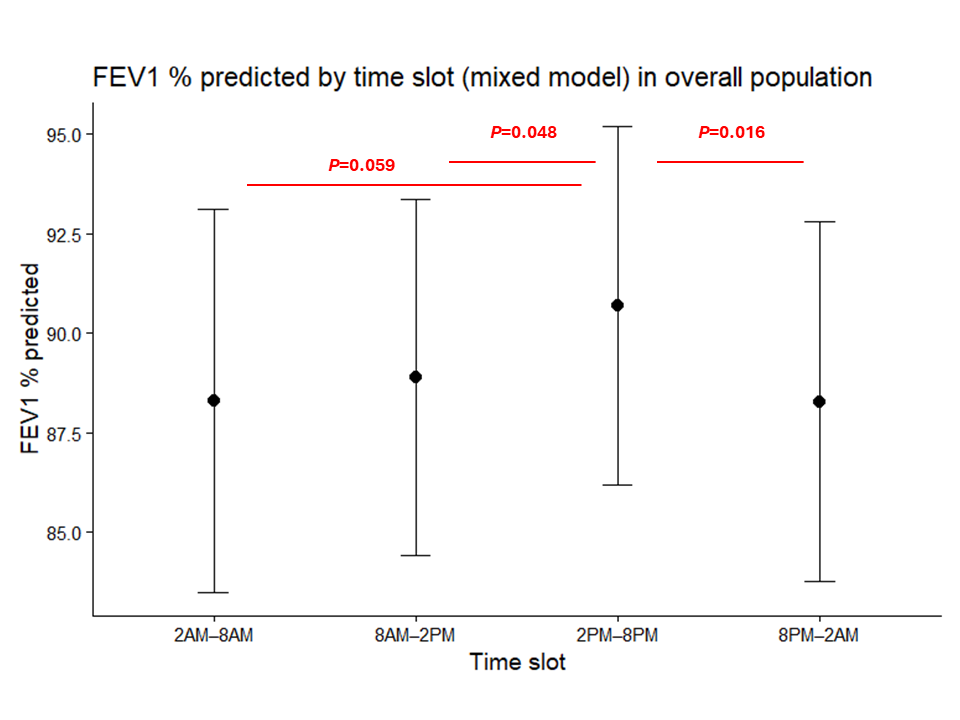

***Figure E20***. Using mixed effect models, FEV_1_ % predicted was higher between 2PM and 8PM but only observed in asthmatics.

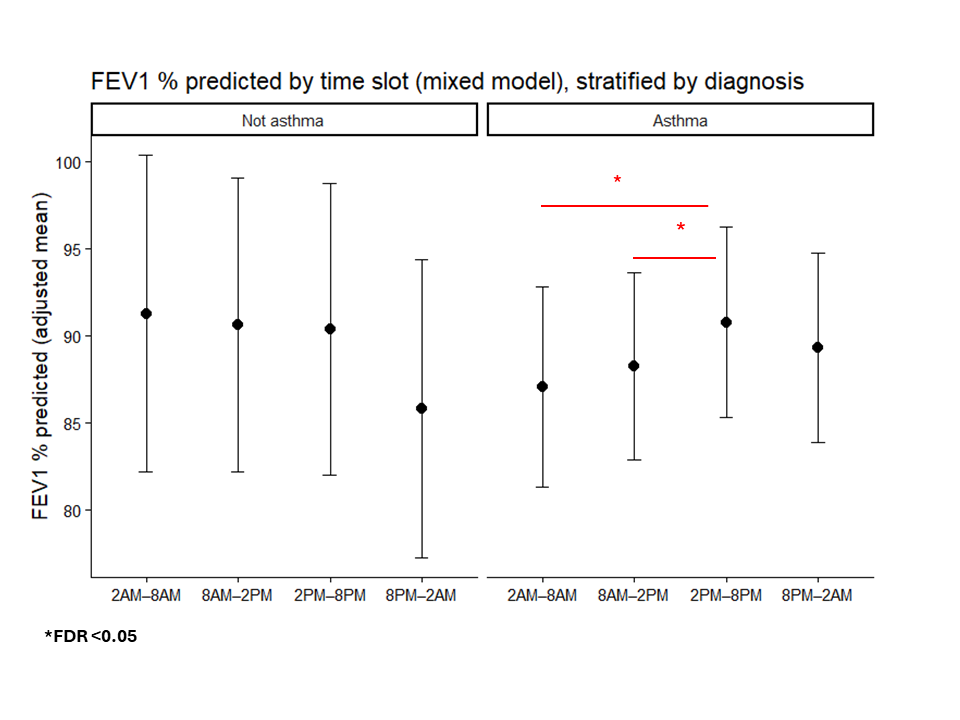

***Figure E21.*** Using mixed effect models, FEV1/FVC was higher between 2PM and 8PM in the overall population.

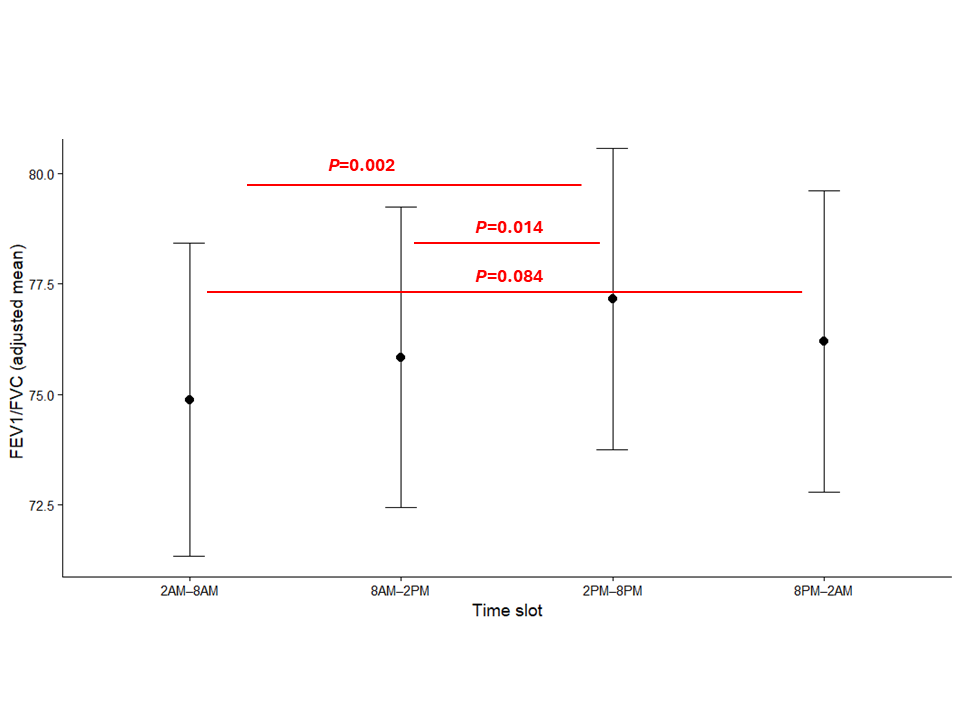

***Figure E22.*** Using mixed effect models, FEV1/FVC was higher between 2PM and 8PM in in asthmatics; this was not observed in those without asthma.

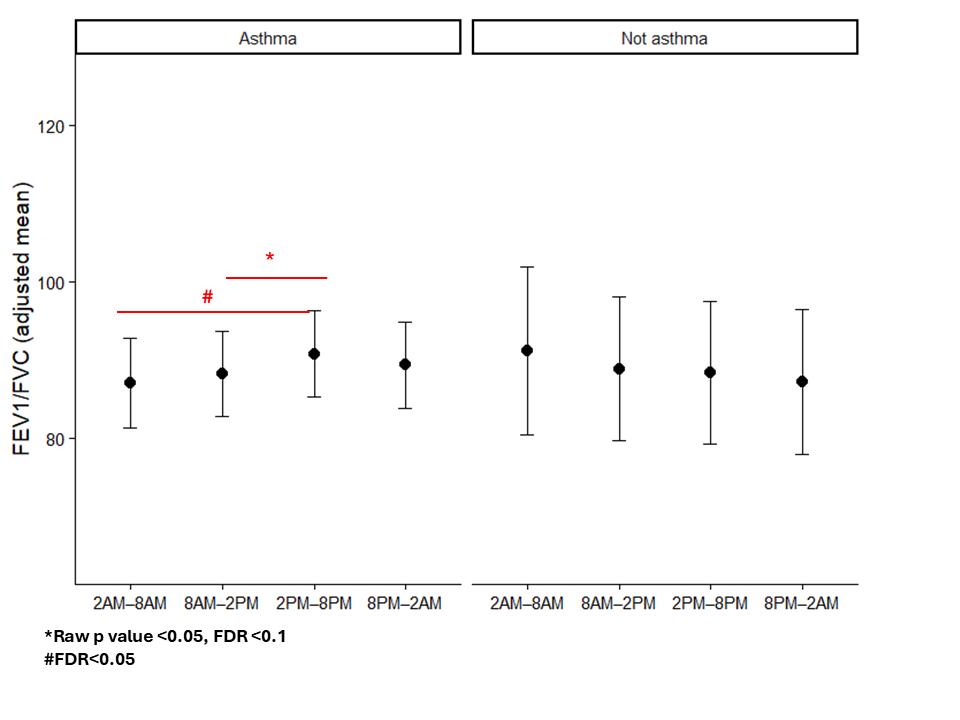

***Figure E23:*** Differences in the putative home spirometry parameters in asthma and those without.

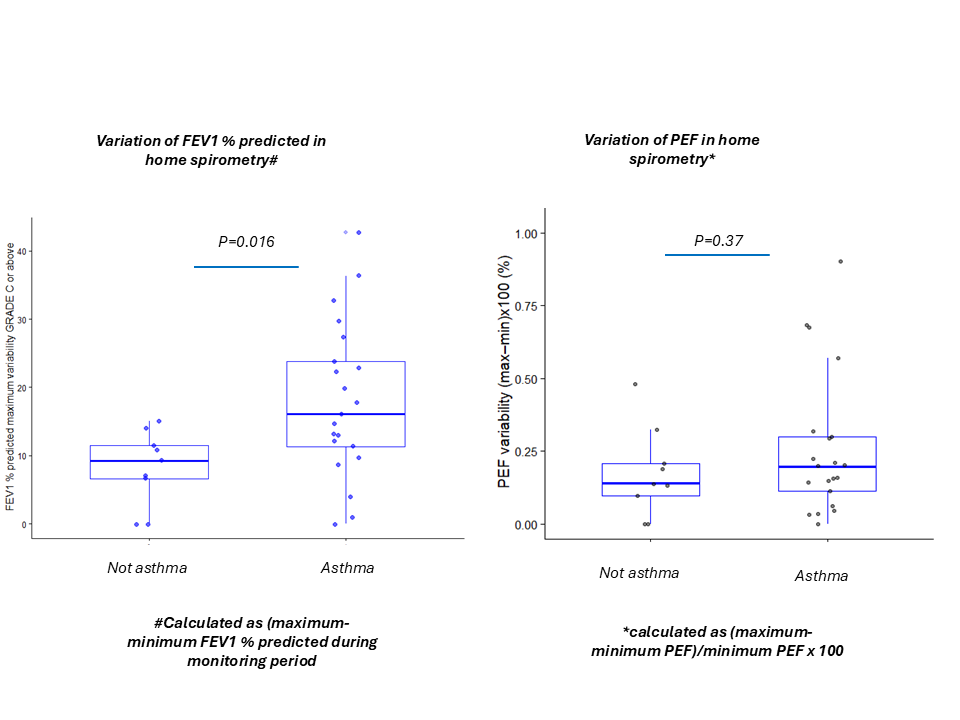

***Table E4.*** *Putative parameters of spirometry using grade A or grad A and B measurements*

| **Potential parameters** | **Number (n)** | **AUC_ROC_** | **Threshold for specificity to be >90%** | **Sensitivity** | **Specificity** |
| --- | --- | --- | --- | --- | --- |
| ***GRADE A measurements only*** | | | | | |
| FEV_1_%Pred_max-min_, (%) | 24 | 0.778 (0.582-0.974) | ≥28% | 11.1 (0-27.8)% | 100 (100-100)% |
| PEF_max-min,_ (%) | 24 | 0.546 (0.307, 0.786) | ≥42% | 22.2 (5.6-57.1)% | 100 (100-100)% |
| ***GRADE A & B measurements only*** | | | | | |
| FEV_1_%Pred_max-min_, (%) | 27 | 0.736 (0.544-0.929) | n/a ≥16% | 50 (30.0-70.1)% | 100 (100-100)% |
| PEF_max-min,_ (%) | 27 | 0.486 (0.23-0.719) | ≥60% |  |  |

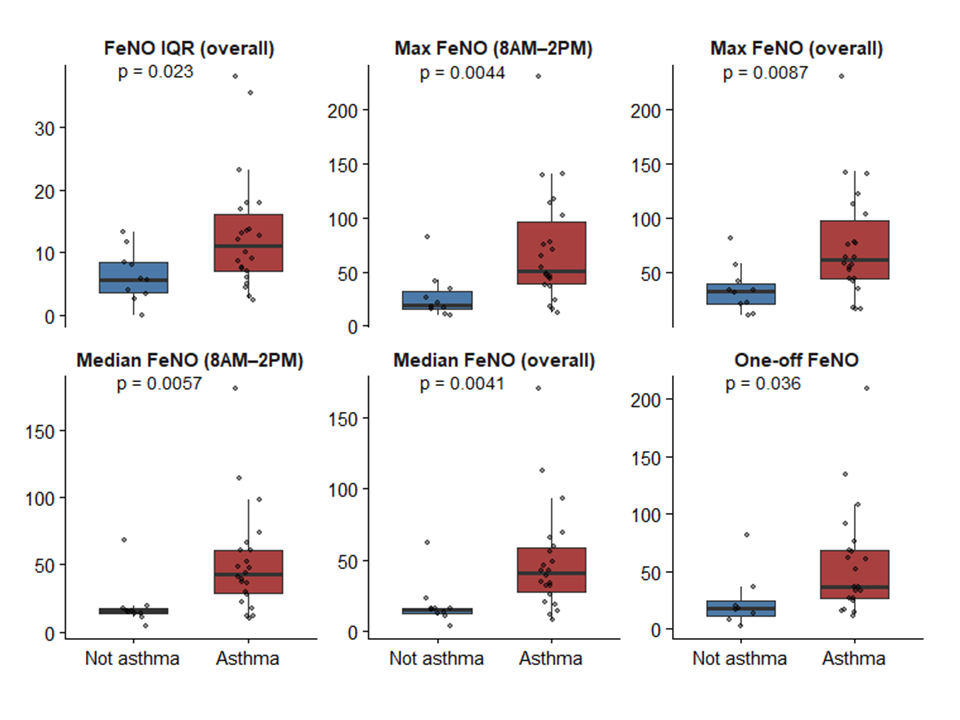
***Figure E24.*** Home FeNO parameters in those with asthma and not asthma.

***Figure E25.*** *Diagnostic efficiency of home FeNO derived parameters.*

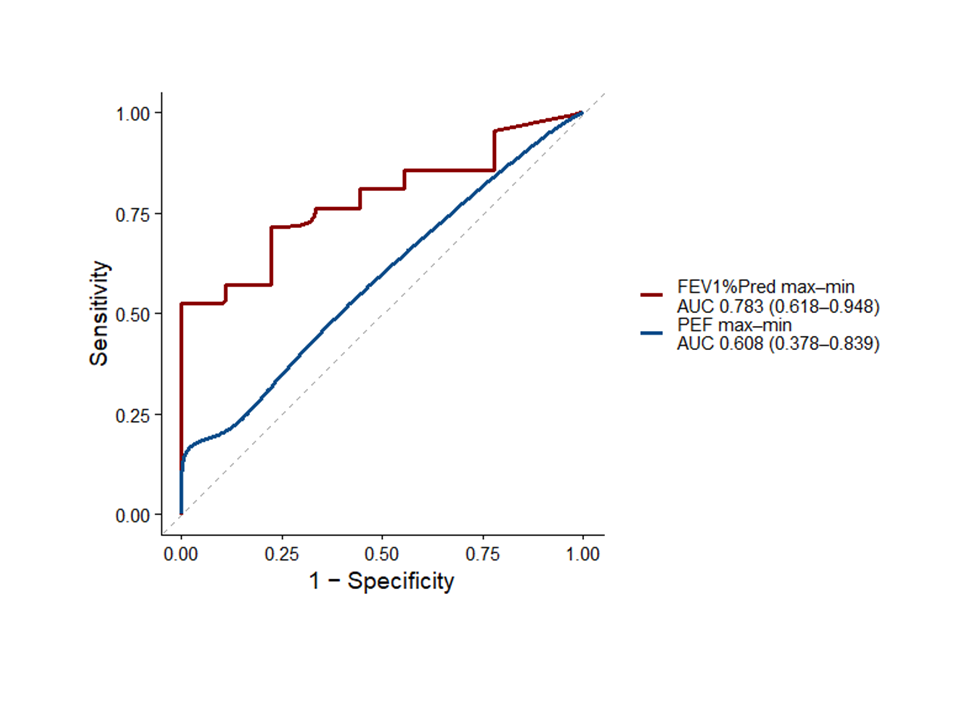

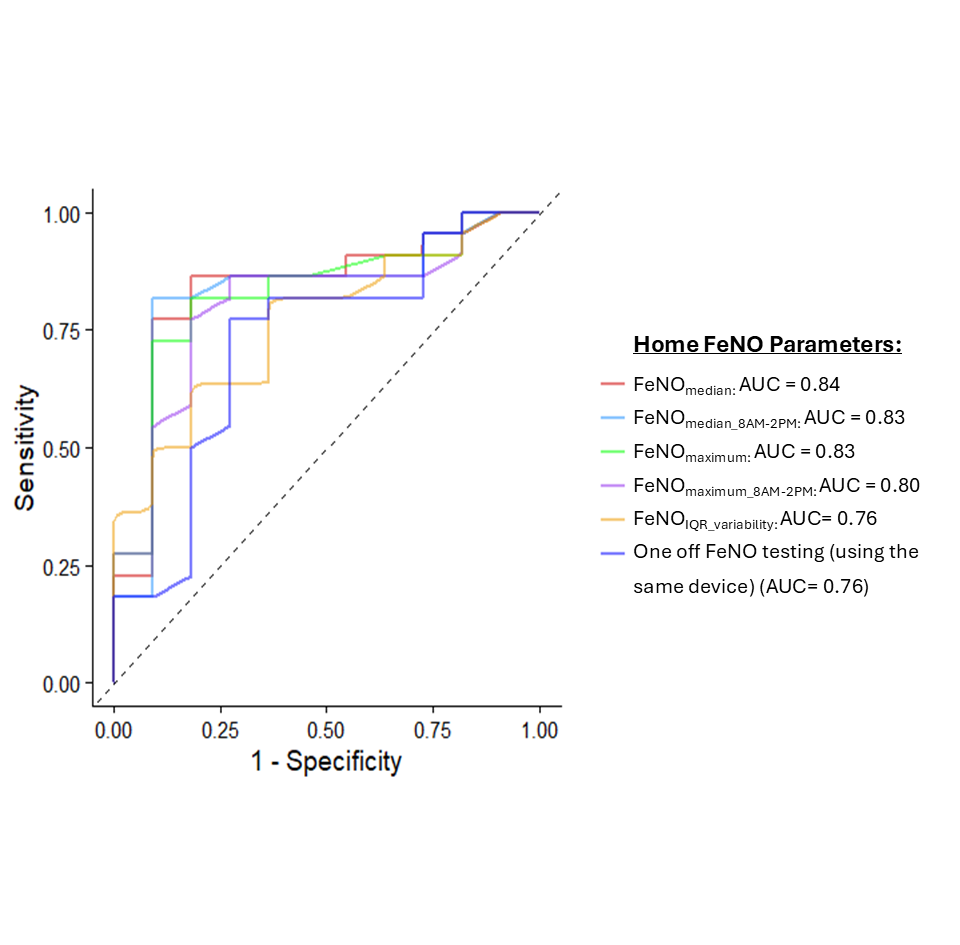

***Figure E26****: Agreement between clinic-based and home FeNO measurements (using NOBreath device) was assessed using Bland–Altman analysis. A mean bias of 6.3 ppb was observed, with clinic-based measurements higher on average than home measurements. The 95% limits of agreement ranged from -30 to +42 ppb, indicating moderate agreement between methods.*
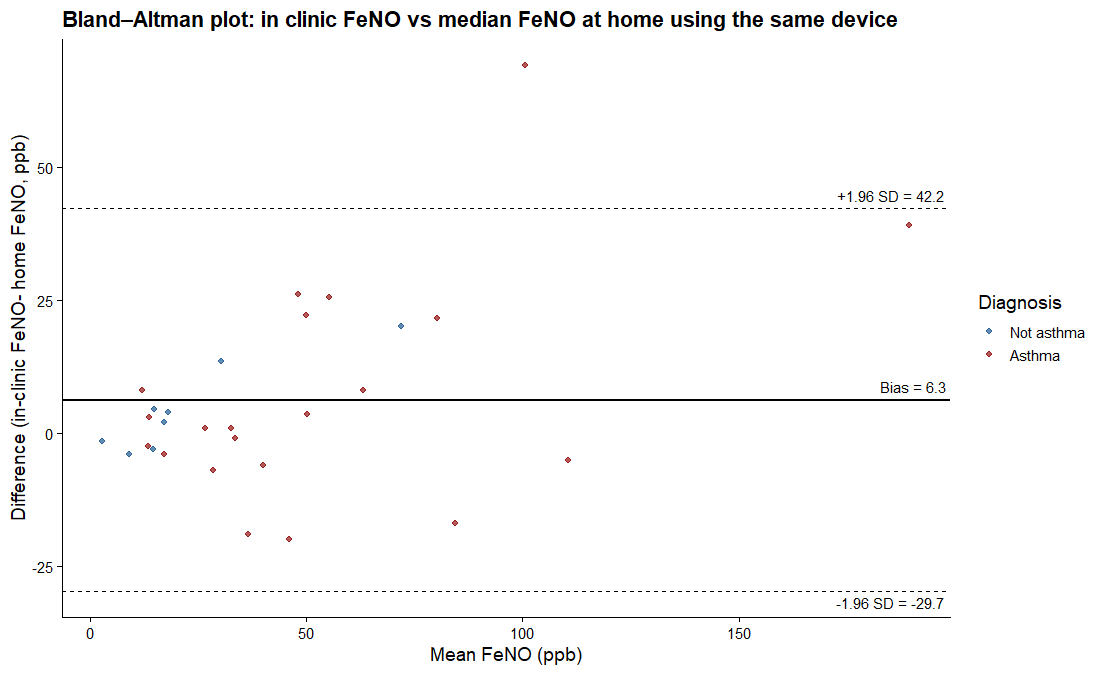

***Table E5.*** *Clinic-based FeNO alternative cutoffs in the current cohort*

| **Potential parameters** | **Number (n)** | **Cutoffs (ppb)** | **Sensitivity** | **Specificity** |
| --- | --- | --- | --- | --- |
| FeNO (NIOX)^§^ | 38 | 25 | 76.0 (60-92)% | 84.6 (61.5-100)% |
| FeNO NOBreath in clinic^Ω^ | 32 | 25 | 72.7 (54.6-90.1)% | 80.0 (50.0-100.0)% |

***§****Considered by Expert Panel to determine reference standard.*

**Ω***Blinded to Expert Panel when reference standard was decided*

**Figure E27.** Application of the integrated home-testing diagnostic pathway in participants with a confirmed asthma outcome. Diagnostic accuracy analysis demonstrated that both proposed integrated home-testing pathways outperformed the BTS/NICE/SIGN 2024 pathway, while substantially reducing the requirement for bronchial challenge testing within this subgroup.

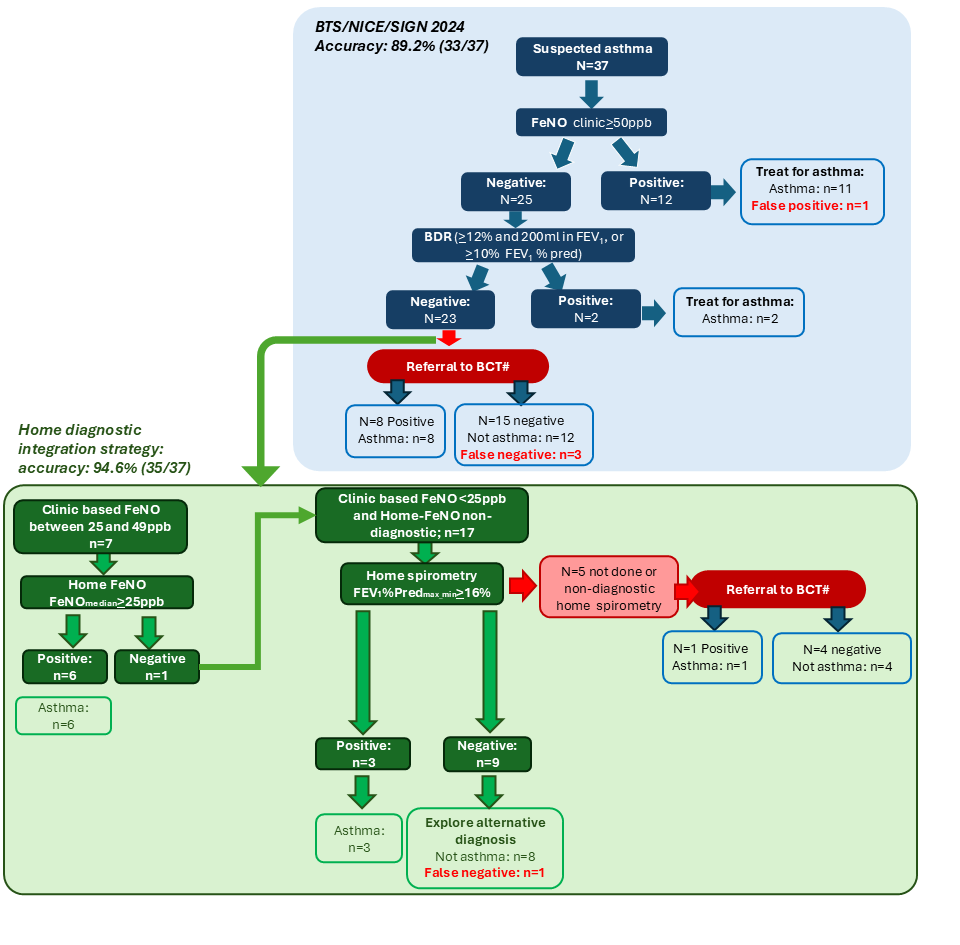

**Table E6. Early economic scoping of home testing:** Early economic scoping using NICE unit costs² indicated that home testing would be cost-neutral on diagnostic costs alone if it reduced bronchial challenge testing (BCT) by 25%, assuming a cost of £45 per patient. This estimate assumes that reusable devices are loaned and returned following testing periods, with the per-patient cost covering mouthpieces and other consumables. In the current feasibility cohort (n=51), the home pathway was estimated to reduce the proportion requiring BCT from approximately 65% to 23.5%, representing a relative reduction of 63.8%. These preliminary estimates exclude potential additional savings from avoided outpatient attendances, unscheduled healthcare use while awaiting diagnostic testing, and secondary-care review before BCT. An exploratory national extrapolation (UK) suggested that this reduction could potentially avoid approximately £5-13 million in BCT costs annually. However, this estimate is highly uncertain and depends on the configuration and costs of current standard care, which are heterogeneous, as well as pathway uptake, device and consumable costs, adherence to home testing, the reduction achieved in BCT referrals, and the costs of delivering home testing at scale.

| **Home-test cost basis** | **Cost per patient** | **BCT reduction required to break even** |
| --- | --- | --- |
| NICE marginal cost (consumables only) | £45 | 25% |
| 1.5 × marginal cost | £68 | 38% |
| 2 × marginal cost | £90 | 50% |
| 2.5 × marginal cost | £113 | 63% |
| 3 × marginal cost | £135 | 75% |

*Break-even estimates include diagnostic costs only and were calculated as the home-testing cost divided by the NICE unit cost of a bronchial challenge test (£179.49). Source: NICE NG245 published unit costs (Tables 29-30)^2^, applied to the current feasibility data.*

2. Asthma: diagnosis, monitoring and chronic asthma management (update). Cost-utility analysis: Most cost-effective sequence or combination of tets to diagnose asthma. BTS/NICE/SIGN collaborative guideline NG 245: Economic analysis report. November 2024
